# Magnetoencephalography-based interpretable automated differential diagnosis in neurodegenerative diseases

**DOI:** 10.1101/2024.06.17.24309023

**Authors:** D Klepachevskyi, A Romano, B Aristimunha, M Angiolelli, F Trojsi, S Bonavita, G Sorrentino, V Andreone, R Minino, Troisi Lopez E, A Polverino, V Jirsa, A Saudargienė, M.-C. Corsi, P Sorrentino

## Abstract

Automating the diagnostic process steps has been of interest for research grounds and to help manage the healthcare systems. Improved classification accuracies, provided by ever more sophisticated algorithms, were mirrored by the loss of interpretability on the criteria for achieving accuracy. In other words, the mechanisms responsible for generating the distinguishing features are typically not investigated. Furthermore, the vast majority of the classification studies focus on the classification of one disease as opposed to matched controls. While this scenario has internal validity, concerning the appropriateness toward answering scientific questions, it does not have external validity. In other words, differentiating multiple diseases at once is a classification problem closer to many real-world scenarios. In this work, we test the hypothesis that specific data features hold most of the discriminative power across multiple neurodegenerative diseases. Furthermore, we perform an explorative analysis to compare metrics based on different assumptions (concerning the underlying mechanisms). To test this hypothesis, we leverage a large Magnetoencephalography dataset (N=109) merging four cohorts, recorded in the same clinical setting, of patients affected by multiple sclerosis, amyotrophic lateral sclerosis, Parkinson’s disease, and mild cognitive impairment. Our results show that it is possible to reach a balanced accuracy of 67,1% (chance level = 35%), based on a small set of (non-disease specific) features. We show that edge metrics (defined as statistical dependencies between pairs of brain signals) perform better than nodal metrics (considering region while disregarding the interactions. Moreover, phase-based metrics slightly outperform amplitude-based metrics. In conclusion, our work shows that a small set of phase-based connectivity metrics applied to MEG data successfully distinguishes across multiple neurological diseases.

## Introduction

In the last twenty-five years, the widespread availability of large-scale brain functional data in health and disease has brought great hope toward discovering the mechanisms underpinning brain diseases and the appearance of neurological symptoms. Cognitive functions emerge from the coordinated interactions among brain regions, manifesting as statistical dependencies among the corresponding brain signals. The overall statistical dependencies between all pairs of signals are often referred to as “functional connectivity” (Friston, 1994). Functional connectivity (FC) is subject-specific and allows subject identification (Finn & Rosenberg, 2021), is altered during the execution of tasks (Corsi et al., 2020), in different environmental conditions (Shine et al., 2016), as well as in neurological diseases (Sorrentino et al., n.d., 2018, 2019). The commonest and most straightforward approach to assessing statistical dependencies has been using descriptive metrics (e.g., Pearson’s correlation). This approach has no underlying assumptions concerning the mechanism underlying the observed statistical dependencies. Other techniques take a more mechanisms-driven approach. As an example, the hypothesis of communication through coherence posits that the occurrence of communication between regions might occur via more or less synchronization (Fries, 2015). Then, metrics such as the Phase Locking value (PLV) were developed to quantify communication via the synchronization between brain signals (such as electroencephalography-EEG and magnetoencephalography-MEG) (Bastos & Schoffelen, 2016). These metrics have been classically used to characterize multiple neurodegenerative diseases (Stam, 2010). More recently, it was shown that large-scale brain activity is far from stationary, and instead, it is characterized by aperiodic, scale-free bursts of activity (Haldeman & Beggs, 2005; Shriki et al., 2013; Tagliazucchi et al., 2012). Then, borrowing from statistical mechanics, the dependencies among brain regions were understood as the presence of scale-free bursts of activities, named *“neuronal avalanches*”, that describe the presence of aperiodic, non-linear bursts of activities spreading brain regions. Intriguingly, in several neurological diseases (such as Parkinson’s disease, Amyotrophic lateral sclerosis, and Mild Cognitive Impairment), brain dynamics spread differently with respect to healthy controls (Polverino et al., 2024; Romano et al., 2023; Sorrentino et al., 2019), and, more importantly, changes in the way aperiodic waves spread proved to be strongly predictive of individual clinical disability (Polverino et al., 2024; Romano et al., 2023).

Despite extensive efforts, there has been a lack of replicability of the studies, regardless of the particular technique adopted to estimate functional connectivity (Kelly & Hoptman, 2022). In other words, the measurements and metrics devised to this day might fail to optimally capture disease-relevant mechanisms comprehensively. As a consequence, automatic classification among multiple neurological diseases cannot be achieved with high accuracy based on functional data alone.

In this paper, we take a different approach and start from the assumption that the way pathophysiological processes spread across the brain has some aspects to it that are specific to a given disease and can be best measured in a set of features that are (spatially) shared among multiple diseases. As a direct consequence, functional connectivity should show specific elements that distinguish various diseases. Hence, the first hypothesis of our study is that it is possible to identify a (small) set of features that can classify multiple neurological diseases.

To test our hypothesis, we leveraged a vast cohort of source-reconstructed MEG data from patients affected by mild cognitive impairment (MCI), amyotrophic lateral sclerosis (ALS), Parkinson’s disease (PD), and Multiple Sclerosis (MS).

First, we compared the classification performance of four FC metrics that capture different properties of the signals (AEC, PLV, Pearson’s correlation coefficient, and ATM) associated with a four-class problem (i.e. MCI, PD, MS, and ALS). We considered the features that can differentiate the considered neurological diseases for each FC metric taken separately.

Furthermore, we compared nodal and edge metrics, under the hypothesis that edges, which more directly represented the interactions among brain regions, would outperform local (i.e.) metrics. We compared the classification performance using three different machine learning algorithms (i.e., XGBoost, Support Vector Machine (SVM), and Linear Discriminant Analysis (LDA), to demonstrate that the performance of a given feature-set is algorithm-independent. Finally, for each FC metric, we identified the most informative features used by the classifier, under the hypothesis that such relevant features were linked to the neurophysiology of the considered neurological diseases. Such a study would make the classification results more interpretable and would enable us to identify clusters of brain interactions sensitive to the neurophysiological mechanisms associated with the considered diseases.

The purpose of this work is to explore a diverse set of connectivity metrics to propose an interpretable automated pipeline for differentiated diagnosis of neurodegenerative diseases.

## 1. Materials and Methods

### 2.1 Participants

One hundred nine patients with different neurological diseases (ALS, MCI, PD, MS) were recruited from Hermitage Capodimonte Clinic in Naples (Polverino et al., 2022; Romano et al., 2023; Sorrentino et al., 2019, 2022). Specifically, Thirty-two MCI patients (18 males and 14 females; mean age 71.31; SD ± 6.83; mean education 10.54; SD ± 4.33) were recruited from the Center of Cognitive and Memory Disorders of the Hermitage Capodimonte Clinic in Naples, Italy. The MCI diagnosis was done according to the National Institute on Ageing-Alzheimer’s Association criteria (Albert et al., 2011). Thirty-nine ALS patients (29 males and 10 females; mean age 59.63; SD ± 12.87; mean education 10.38 years SD ± 4.3) were selected in collaboration with the ALS Center of the First Division of Neurology of the University of Campania “Luigi Vanvitelli” (Naples, Italy). The ALS diagnosis was performed according to the El-Escorial criteria (Brooks, 1994). Twenty patients (14 males and 6 females; mean age 64.5; SD ± 12.18; mean education 11 years SD ± 3.9) with a confirmed diagnosis of Parkinson’s disease according to the United Kingdom Parkinson’s Disease Brain Bank criteria (Gibb & Lees, 1988) were recruited in collaboration with the Movement Disorder Unit of Cardarelli hospital in Naples. Finally, eighteen patients (6 males and 12 females; mean age 45.05; SD ± 9.92; mean education 14-11 years SD ± 4.89) with Multiple Sclerosis were recruited in collaboration with University of Campania Luigi Vanvitelli. The diagnosis was performed following the 2017 revision of the McDonald criteria (Thompson et al., 2018). Each participant underwent a specific motor and/or neuropsychological evaluation according to the clinical characteristics of each disease. A complete summary of the cohort description is available in Table 1. The study protocol was approved by the ‘‘Comitato Etico Campania Centro’’ (Prot.n.93C.E./Reg. n.14-17OSS) and all participants provided written informed consent in accordance with the Declaration of Helsinki.

**Table 1:** Demographic features of the cohort: m: males; f: females; SD: Standard Deviation.

| *Type of disease* | *Number of participants (109)* | *Age (mean ± SD)* | *Years of education (mean ± SD)* | *Gender (ratio)* |
| --- | --- | --- | --- | --- |
| *Mild Cognitive Impairment (MCI)* | 32 | 71.31<br>(SD ± 6.83) | 10.54<br>(SD ± 4.33) | 18 m / 14 f |
| *Multiple Sclerosis (MS)* | 18 | 45.05<br>(SD ± 9.92) | 14.11<br>(SD ± 4.89) | 6m /12 f |
| *Parkinson's Disease (PD)* | 20 | 64.5<br>(SD ± 12.18) | 11<br>(SD ± 3.9) | 14 m / 6 f |
| *Amyotrophic Lateral Sclerosis (ALS)* | 39 | 59.63<br>(SD ± 12.87) | 10.38<br>(SD ± 4.3) | 29 m/10 f |

### 2.2 MEG and MRI acquisition, pre-processing, and source reconstruction

MEG and MRI acquisition, preprocessing, and source reconstruction were performed similarly to previous studies (Cipriano et al., 2024, p. 20; Romano et al., 2022). Briefly, all patients underwent an MRI scan using a 3T Biograph mMR tomograph (Siemens HealthcareErlangen, Germany) equipped with a 12 channels head coil. Specifically, 3 dimensional T1-weighted images (gradient-echo sequence inversion recovery prepared fast spoiled gradient recalled-echo, time repetition = 6,988 ms, inversion time = 1,100 ms, echo time = 3.9 ms, flip angle = 10, voxel size = 1 × 1 × 1.2 mm3) were acquired. The MEG acquisition was performed using a 163-magnetometer system placed in a magnetically shielded room (AtB Biomag UG, Ulm, Germany). Fastrack (Polhemus®) was used to define the position of the head under the helmet and to digitalize the position of four anatomical landmarks (nasion, right, and left preauricular and apex) and four reference coils. Each patient performed two recordings of 3.5 minutes each, with a one-minute break, during a resting state, with eyes closed. Electrocardiographic and electrooculographic signals were recorded to remove physiological artifacts. Data were acquired with a sampling frequency of 1024 Hz. A Principal component analysis (PCA) was used to reduce the environmental noise, and an independent component analysis (ICA) was used to remove physiological artifacts (namely ocular and cardiac artifacts). Finally, to obtain the source-reconstructed time series of the patients, according to the Automated Anatomical Labeling (AAL) atlas, we used a beamformer algorithm and the volume conduction model proposed by Nolte (Nolte, 2003). The time series were filtered between 0.5 and 48 Hz.

### 2.3 Connectivity Metrics

#### Phase Locking Value (PLV)

The PLV measures the phase synchronization between two narrowband signals, and it is computed as: (Lachaux et al., 1999).

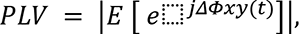

where ΔΦ_xy_*(t)* represents the difference between Φ_x_(*t*) - ΔΦ_y_*(t), [E ]* is the statistical expectation, and ΔΦ_x, y_*(t)* are the instantaneous phases of the analytical signals.

#### Correlation Coefficient (CC)

We computed Pearson’s correlation coefficient to estimate the pairwise synchronization between signals of different brain regions.

#### Avalanche Transition Matrix (ATM)

The ATM describes the probability that after the activation of region *i* at the time *t*, the region *j* will be active at the time t +δ (Sorrentino et al., 2021). The ATMs are computed starting from neuronal avalanches, which are defined as events that start when at least one region is above the threshold and end when all the regions return to their baseline activity. Hence, there is one ATM for each avalanche. More specifically, the ATM contains, in the ijth position, the probability that region j is active at time t+1 given that region t is active at time t.ATMs were then averaged element-wise over all the avalanches for a subject, and finally symmetrized.

#### Amplitude Envelope Correlation (AEC)

The amplitude envelope is used to estimate the statistical interdependencies between brain regions. It is computed as the correlation coefficient between the analytical amplitude of two signals. High values of amplitude correlation between the envelopes indicate that two brain regions display a coordinated behavior (Brookes et al., 2011, 2012).

#### Nodal analysis

Each of the connectivity metrics yields an adjacency matrix. We have compared directly a subset of the entries of the matrices (see section 3.2), that is “edge-metrics” or nodal metrics. Three different edge-specific metrics were used: betweenness centrality, eigenvector centrality, and the degree. Betweenness centrality is a centrality measure that is equal to the number of the shortest paths passing through a given node. Another centrality measure is eigenvector centrality, which determines a node’s relative importance within a network. Lastly, the degree of a node is the sum of the weights of the edges incident upon the node.

### 2.4 Classification Algorithms

To evaluate the discriminative ability of different feature sets (PLV, CC, ATM, AEC) and compare them with each other, we applied three different Machine Learning (ML) algorithms. Balanced accuracy was used as an evaluation metric, since we have imbalanced classes. ML algorithms include Linear Discriminant Analysis (LDA), Support Vector Machines (SVM), and Extreme Gradient Boosting (XGBoost). The general modelling workflow is summarized in Fig. 1

**Figure 1.**
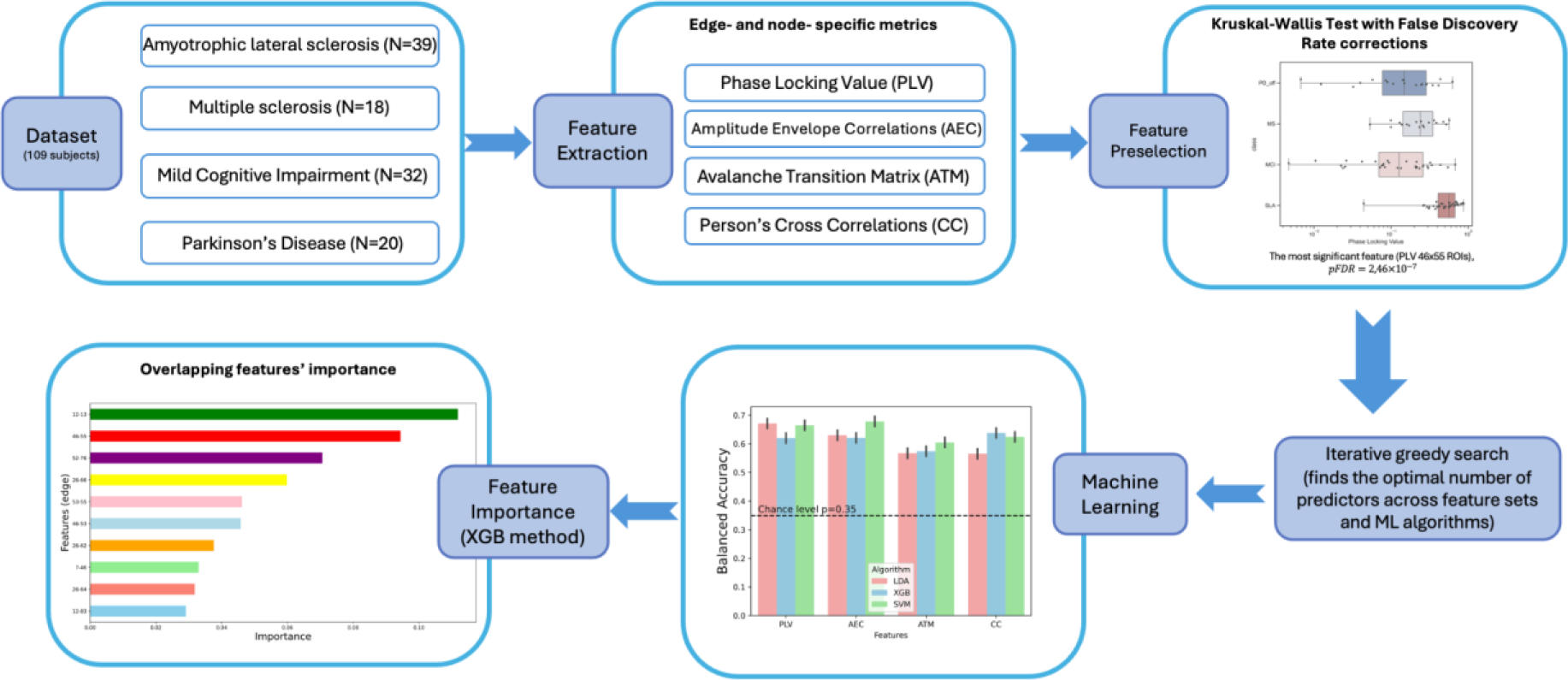
The general workflow of modeling.

#### Linear Discriminant Analysis

LDA is a widely used approach for solving multi-class classification problems. The algorithm separates multiple classes (in our study −4 classes) with multiple features through a data dimensionality reduction approach. LDA aims to find a hyperplane that best separates the classes while minimizing the overlap within each class. Related work has revealed that LDA performs well with multiclass diagnosis problems (Lin et al., 2021).

#### Support Vector Machines

SVM is another widely used technique for solving supervised tasks with multiple classes. Several studies identified SVM as an outstanding algorithm for solving tasks with multiple classes (Maqsood et al., 2022). SVM performs complex data transformations (according to the selected kernel function) and maximizes the separation boundaries between the data points depending on the classes.

#### Extreme Gradient Boosting

Recent studies showed that XGBoost is a state-of-the-art tree-based machine learning model that outperforms many other algorithms, including deep learning models (Grinsztajn et al., 2022). Moreover, the XGBoost algorithm provides an assessment of the relative importance of individual predictors, which allows us to interpret our findings (Manju et al., n.d.)

XGBoost is an ensemble method that builds a predictive model by combining predictions of multiple individual decision trees. It uses weak learner trees, these are decision trees with a single split, called decision stumps. The algorithm works by sequentially adding weak learners to the ensemble, with each new learner focusing on correcting the errors made by the previous one.

XGBoost is known for its high accuracy and has been shown to outperform other machine learning algorithms in many predictive modeling tasks. In addition, it is highly scalable and can handle large datasets.

### 2.5 Statistical Analysis

#### Kruskal-Wallis test

For each connectivity metric taken separately, to identify the most statistically significant different features among the four groups (PD, MCI, SLA, MS) to be considered for the classification, we used the Kruskal-Wallis test. Since the brain is a non-linear dynamic system, we relied on a non-parametric statistical test, checking the null hypothesis that two or more independent groups were drawn from the same underlying distribution. The same approach was used for both edge and nodal metrics.

#### Multiple comparison correction

Since we have numerous features to be considered for a given FC metric, we used the false discovery rate to correct for inflated significance. The False Discovery Rate (FDR) is used to control the expected proportion of false positives. The FDR is the expected ratio of the number of false positive classifications, or false “discoveries”, to the total number of positive classifications (rejections of the null hypothesis). The p-values of the Kruskal-Wallis test were corrected accordingly. Finally, we sorted the features according to the corrected p-values in ascending order.

#### Spearman Correlation

We used Spearman correlation to evaluate the correlation between the features’ ranks. Spearman’s rank correlation coefficient is a non-parametric measure of statistical dependence between two variables. This way, we evaluated the relation between the ranks of the nodal or edge features across different FC metrics (PLV, AEC, ATM).

#### Repeated Stratified K-fold splits

To get valid results and avoid overfitting, we applied Repeated Stratified K-fold cross-validation, which repeats k-folds n times with different randomization for each repetition (J.-H. Kim, 2009). Then, for each fold, we have pooled our results across multiple randomization. First, our whole dataset was split into two parts. For the first part, we use Stratified K-folds cross-validation to tune hyperparameters and find an optimal set that gives the best result. After tuning the hyperparameters on the first part of the dataset, then we used Stratified K-fold cross-validation 10 times for the second part. Then, accuracies obtained by each set are averaged. This way, we prevent data leakage, and it helps to get a more robust estimation of the accuracy by averaging over all repetitions and all folds. We used 10 repetitions of 10-fold cross-validations, and therefore we ensure that our evaluation is not affected by the specific choice of the validation set.

### 2.7 Evaluation metrics

#### Balanced accuracy

We used balanced accuracy as an evaluation metric for the classification algorithms since our dataset is imbalanced (NALS=39, NMCI=32, NPD=20, NMS=18). The balanced accuracy is calculated by taking the average of the recalls obtained in each class (Thölke et al., 2023).

#### Recall

Recall is an evaluation metric that measures how often a classification algorithm correctly identifies positive instances among all the actual positive samples in the dataset.

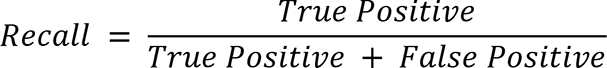

#### Receiver Operating Characteristic (ROC) curve

ROC curve is a graph that displays the performance of a binary classification algorithm of predicting a positive class at all possible thresholds. The lower the classification threshold, the more observations are successfully classified. ROC curve uses False Positive Rate on the x-axis and True Positive Rate on its y-axis.

The area under the ROC curve (AUC) is an evaluation measure that measures the area underneath the ROC curve, and its maximum possible value equals one. In this manuscript, we compute the ROC curve for each class separately.

#### Confusion Matrix

A confusion matrix is an N x N matrix, where N is the number of classes. It has the true labels on the rows and the predicted labels on the second axis. This way, a confusion matrix shows how many times each class was classified correctly and also how often it was misclassified (and how).

We used a confusion matrix for 4 classes, therefore we have a 4 x 4 matrix, where *TP_i_* represents the observations that were correctly classified for class *i*, and *E_ij_* represents where true class *j* was misclassified with predicted class *i*. After that, we took the relative percentages across columns to see the whole picture in percentages, therefore each column’s values will sum up to 100%. This is done by dividing each element of each column by the sum of all elements of that column and multiplying by 100. For example, for column 4 and its third element, it is done as follows:

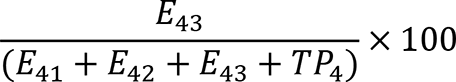

#### Code availability

The code used to perform the analysis of this study is publicly available at https://github.com/dklpp/multiclass_meg_features_analysis

## 2. Results

### 3.1 Kruskal-Wallis Test

Each adjacency matrix obtained from a given FC metric (namely PLV, AEC, ATM, or CC) is a square matrix with the dimension of *n_regions_* × *n_regions_*, where *n_regions_* is equal to 116 regions of interest. All matrices are symmetric and contain ones on the main diagonal., Hence, we take the triangular matrix, excluding the main diagonal elements, leading to 6670 edge-wise features. Given the high dimensionality of the feature space, we identified the most statistically significant different features among the four groups to be considered for the classification.

A non-parametric statistical Kruskal-Wallis test was performed for each feature to compare the four independent groups (PD, SLA, MS, MCI). After applying Kruskal-Wallis Test and False Discovery Rate correction, we found that there were more than 120 statistically significant edge features (pFDR < 0.002) for each of the 4 edge-specific FC metrics. The lowest corrected with FDR p-value p<0.0001 (pFDR = 2,46 × 10^−7^) was obtained with the edge-wise PLV metric between the right frontal superior gyrus and the right postcentral gyrus (see Fig 2)

**Figure 2.**
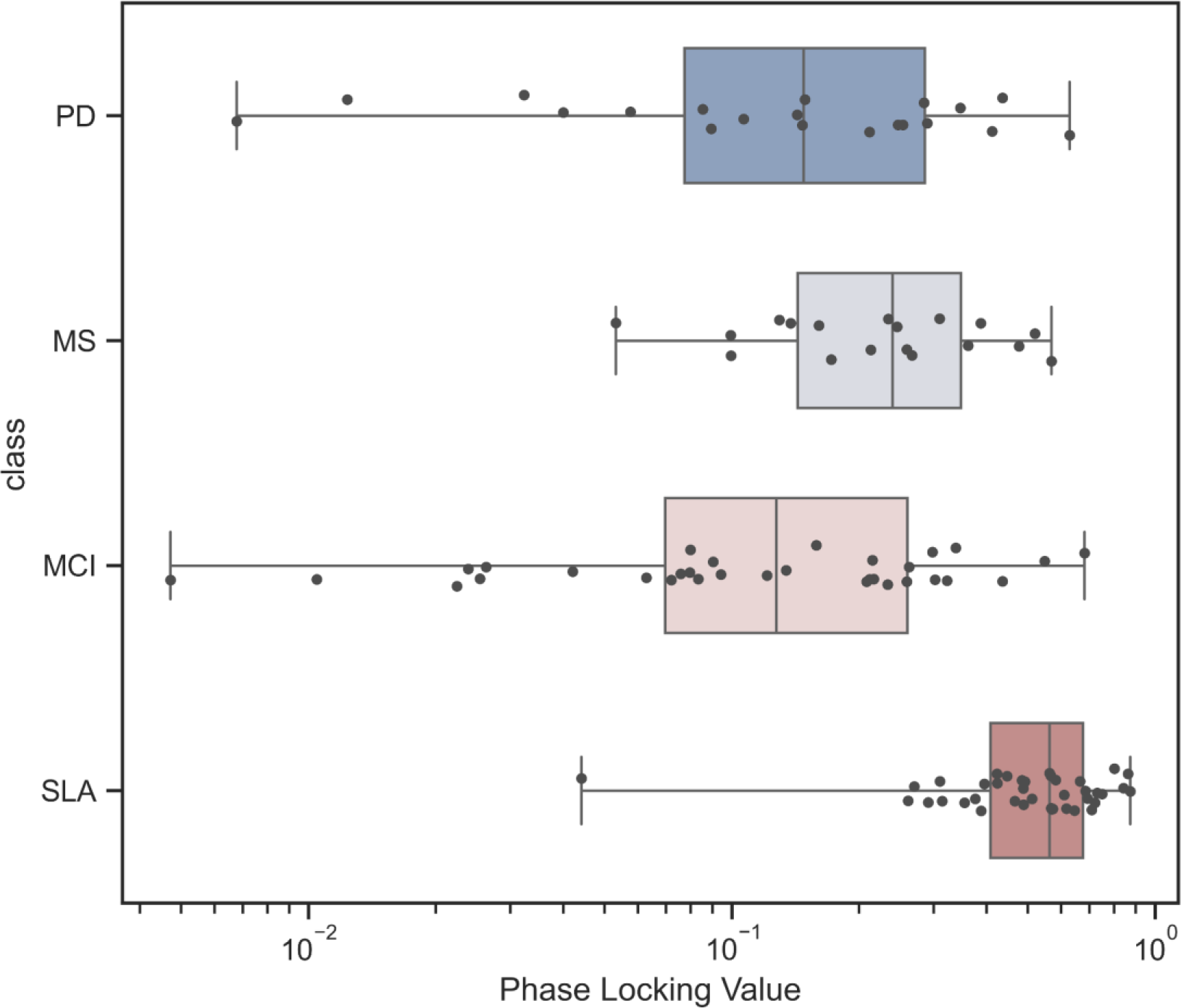
The most significant features’ (PLV values between the right frontal superior gyrus and the right postcentral gyrus) boxplots with the observations for 4 classes with FDR p-value (pFDR = 2,46 x 10^-7^)

### 3.2 Classification algorithms

Based on the significant edges, we have then classified the participants. We used a consecutive iterative search technique, starting with the 15 best features (according to their corrected p-values), and sequentially added features and compared the accuracies, and this procedure was repeated until 39 features were fed to the classifier (Table 2). Stability had been reached at this point, and further increasing the number of features led to a slight worsening of the performance (not shown). Furthermore, given the relatively small size of our sample, we kept a lower number of features to reduce overfitting.

**Table 2:**
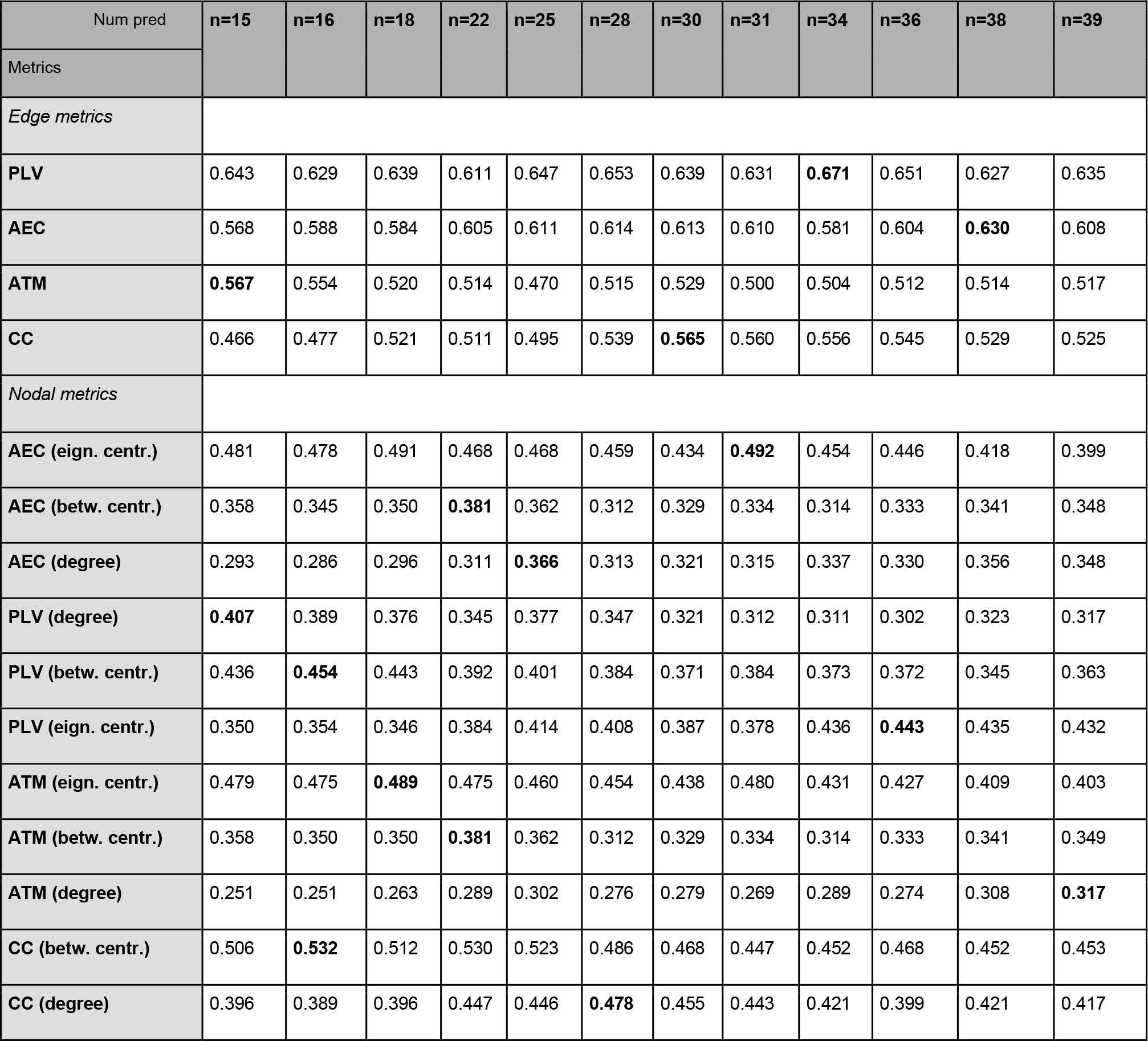
Balanced accuracy for the number of features which contain the best accuracy across different metrics (PLV, AEC, ATM, CC). For visual purposes, it is demonstrated only with the LDA algorithm. The accuracies obtained with the XGBoost and the SVM algorithms are in the supplementary material (see S4 and S5).

| Num pred | n=15 | n=16 | n=18 | n=22 | n=25 | n=28 | n=30 | n=31 | n=34 | n=36 | n=38 | n=39 |
| --- | --- | --- | --- | --- | --- | --- | --- | --- | --- | --- | --- | --- |
| Metrics |  |  |  |  |  |  |  |  |  |  |  |  |
| *Edge metrics* |  |  |  |  |  |  |  |  |  |  |  |  |
| **PLV** | 0.643 | 0.629 | 0.639 | 0.611 | 0.647 | 0.653 | 0.639 | 0.631 | **0.671** | 0.651 | 0.627 | 0.635 |
| **AEC** | 0.568 | 0.588 | 0.584 | 0.605 | 0.611 | 0.614 | 0.613 | 0.610 | 0.581 | 0.604 | **0.630** | 0.608 |
| **ATM** | **0.567** | 0.554 | 0.520 | 0.514 | 0.470 | 0.515 | 0.529 | 0.500 | 0.504 | 0.512 | 0.514 | 0.517 |
| **CC** | 0.466 | 0.477 | 0.521 | 0.511 | 0.495 | 0.539 | **0.565** | 0.560 | 0.556 | 0.545 | 0.529 | 0.525 |
| *Nodal metrics* |  |  |  |  |  |  |  |  |  |  |  |  |
| **AEC (eign. centr.)** | 0.481 | 0.478 | 0.491 | 0.468 | 0.468 | 0.459 | 0.434 | **0.492** | 0.454 | 0.446 | 0.418 | 0.399 |
| **AEC (betw. centr.)** | 0.358 | 0.345 | 0.350 | **0.381** | 0.362 | 0.312 | 0.329 | 0.334 | 0.314 | 0.333 | 0.341 | 0.348 |
| **AEC (degree)** | 0.293 | 0.286 | 0.296 | 0.311 | **0.366** | 0.313 | 0.321 | 0.315 | 0.337 | 0.330 | 0.356 | 0.348 |
| **PLV (degree)** | **0.407** | 0.389 | 0.376 | 0.345 | 0.377 | 0.347 | 0.321 | 0.312 | 0.311 | 0.302 | 0.323 | 0.317 |
| **PLV (betw. centr.)** | 0.436 | **0.454** | 0.443 | 0.392 | 0.401 | 0.384 | 0.371 | 0.384 | 0.373 | 0.372 | 0.345 | 0.363 |
| **PLV (eign. centr.)** | 0.350 | 0.354 | 0.346 | 0.384 | 0.414 | 0.408 | 0.387 | 0.378 | 0.436 | **0.443** | 0.435 | 0.432 |
| **ATM (eign. centr.)** | 0.479 | 0.475 | **0.489** | 0.475 | 0.460 | 0.454 | 0.438 | 0.480 | 0.431 | 0.427 | 0.409 | 0.403 |
| **ATM (betw. centr.)** | 0.358 | 0.350 | 0.350 | **0.381** | 0.362 | 0.312 | 0.329 | 0.334 | 0.314 | 0.333 | 0.341 | 0.349 |
| **ATM (degree)** | 0.251 | 0.251 | 0.263 | 0.289 | 0.302 | 0.276 | 0.279 | 0.269 | 0.289 | 0.274 | 0.308 | **0.317** |
| **CC (betw. centr.)** | 0.506 | **0.532** | 0.512 | 0.530 | 0.523 | 0.486 | 0.468 | 0.447 | 0.452 | 0.468 | 0.452 | 0.453 |
| **CC (degree)** | 0.396 | 0.389 | 0.396 | 0.447 | 0.446 | **0.478** | 0.455 | 0.443 | 0.421 | 0.399 | 0.421 | 0.417 |

We decided to display the balanced accuracies for each FC metric taken separately (both edge-based and node-based metrics) to see a clearer picture of different sets’ performances (Fig 3).

**Figure 3.**
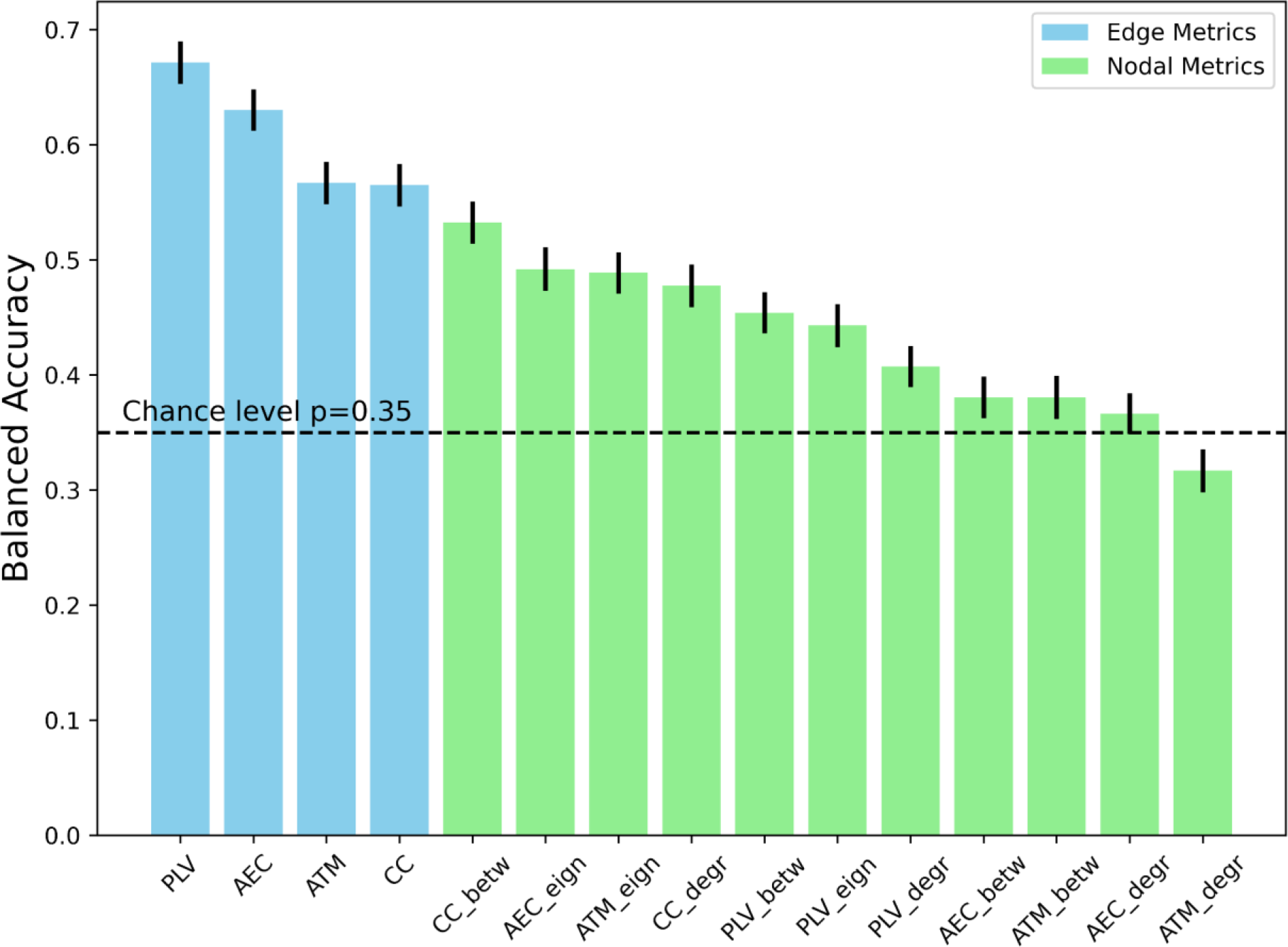
The balanced accuracies for all feature sets with LDA classifier. Each bar plot displays the averaged accuracy with its standard errors. The boxplots for other algorithms are available in the supplementary material (Fig S1-S2)

As shown in Fig3, we observed that all edge metrics consistently outperformed nodal metrics. Consistently, we observed that the standard deviations (over different repetitions of the K-folds) were higher for nodal metrics. Note that the results refer to the best-performing feature selection (i.e. the number of features is not fixed across different metrics).

Finally, we identified the optimal number of features (i.e. the features that showed the lowest corrected p-values and which led to the highest balanced accuracy) for each of the 3 different Machine Learning classification algorithms considered (namely XGBoost, SVM, LDA)(Fig. 3). The exhaustive search algorithm yielded the feature sets (which nodes/ edges) with the best-balanced accuracies for each algorithm across different FC metrics.

For the sake of simplicity, we discuss here the two best-performing FC metrics per classification algorithm (Fig. 4). In the case of the SVM algorithm, the AEC showed a balanced accuracy of 67.8% with a total of 31 top features, the PLV presented a balanced accuracy of 66.5 % with 36 top features. With the XBoost classifier, the CC showed a balanced accuracy of 63.8% with 35 top features), and the PLV presented a balanced accuracy of 62.8%, with 15 top features. Finally, in the case of the LDA classifier, the PLV showed a balanced accuracy of 67.1%, with 34 top features and the AEC presented a performance of 63.0% with 38 top features.

**Figure 4.**
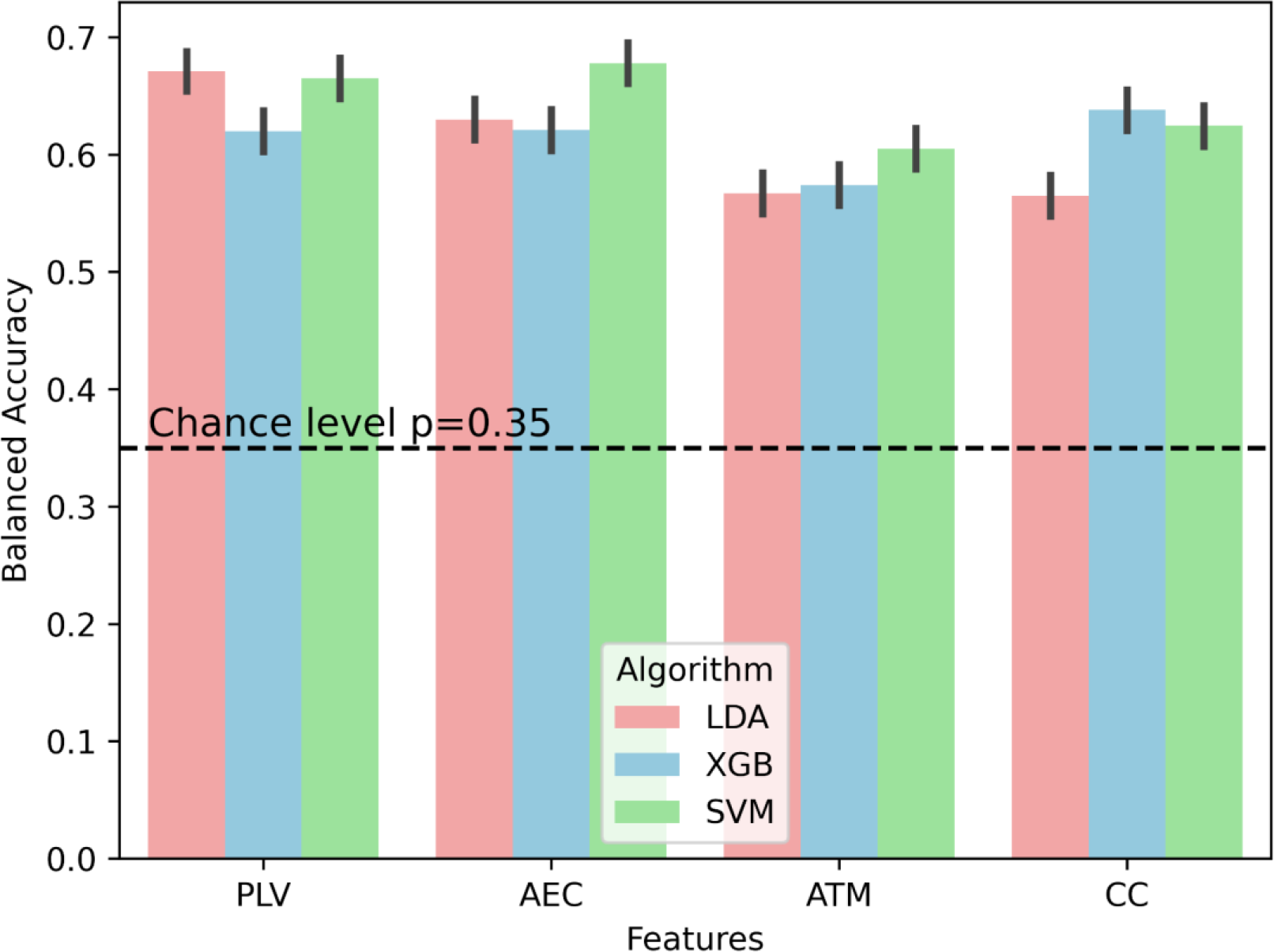
Balanced accuracies for different metrics across 3 Machine Learning algorithms with its standard errors. We observe consistent higher performance of PLV and AEC, in comparison to ATM and CC. The chance level for our dataset equals 35%.

Given the lower performance obtained with the nodal metrics, we shall proceed with the analyses exclusively on the edges.

The chance level for the balanced problem with 4 classes is equal to 25%. However, since we have a dataset with unbalanced classes, in such tasks the chance level is usually assumed to be the probability of predicting the most frequent class label in the target.In our case, SLA, which contains 38 patients out of 108, is the most numerous class. Therefore, the chance level is calculated as follows:

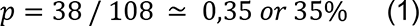

In this task, a more objective evaluation metric is the balanced accuracy. Nevertheless, it is also useful to compare and evaluate overall accuracies. The trend remains the same – i.e. the same edge-specific metrics stay as the top features sets. Still, the accuracies are slightly higher: AEC (73.3%, 28 features), PLV (72.7%, 36 features), ATM (67.5%, 32 features), CC (69.1%, 36 features) for SVM classifier; AEC (68.2%, 38 features), PLV (69.5%, 34 features), ATM (63.2%, 15 features), CC (60.4%, 30 features) for LDA classifier; AEC (68.5%, 26 features), PLV (68.7%, 15 features), ATM (64.2%, 39 features), CC (70.5%, 35 features) for XGBoost classifier.

Since the PLV is the most performant metric, we now focus on the PLV for sensitivity analyses.

### 3.3 ROC curves

After repetitive stratified K-folds, we can estimate probabilities for each class to be correctly predicted (i.e. the probabilities sum up to 1) with different classification algorithms (LDA, SVM, XGB). We applied the One-vs-All technique, where we fix one desired class and all other classes are treated as one class. This way, we can replace our multi classification task to a binary class, and it enables us to build the ROC curves.

We calculated the average ROC curve for each repetition of the 10 folds, and the red curve displays the overall average ROC curve across 10 repetitions. The ROC curves were built for each class separately (Fig 5) which display the trade-off between False Positive Rate on x axis, and True Positive Rate on y axis.

**Figure 5.**
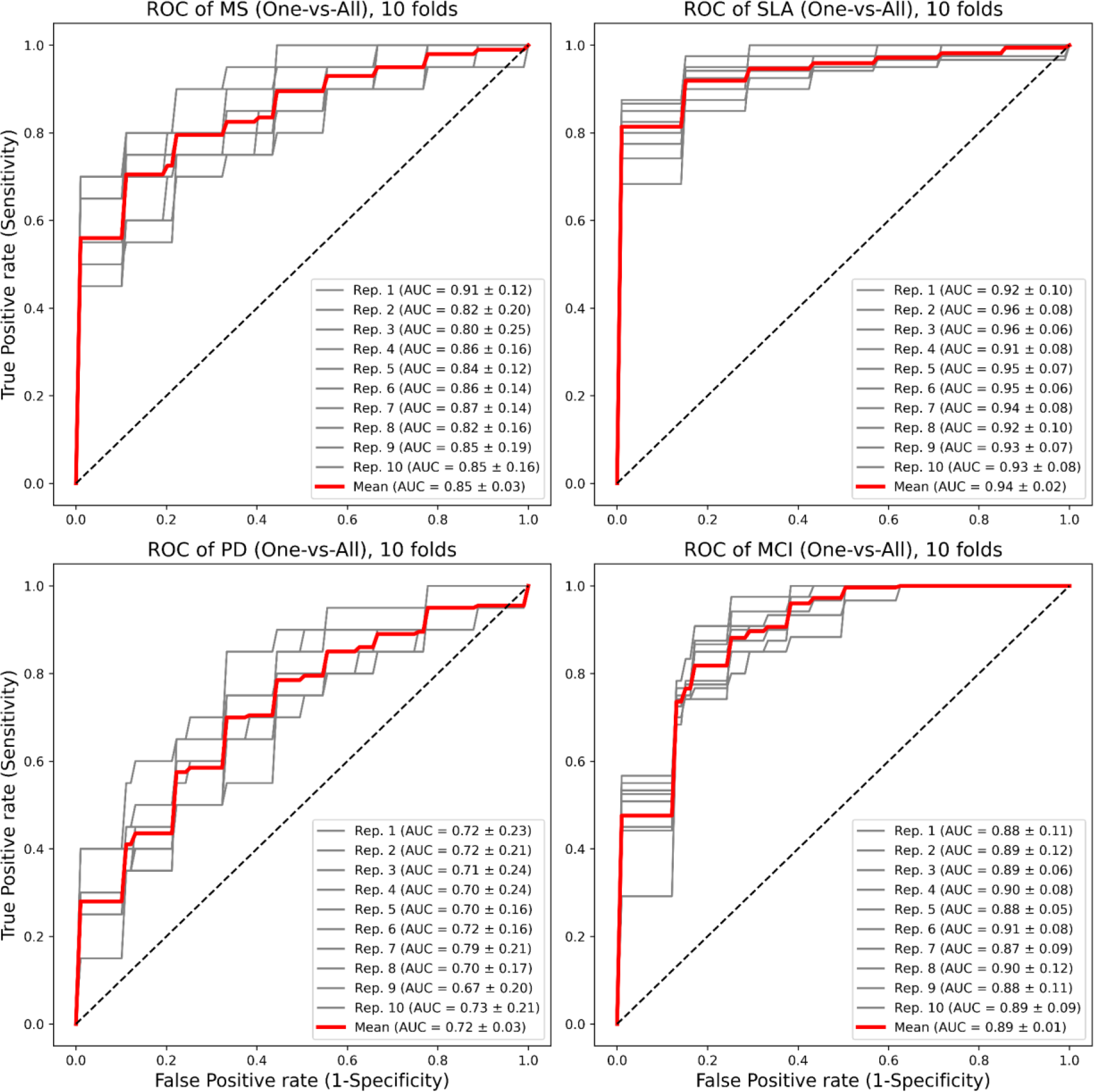
ROC curves for 4 classes (MS, SLA, PD, MCI) for LDA machine learning classifier with overall mean curve and mean curves for each repetition of Stratified K-folds. ROC curves are built with PLV edge-based features with 10 repetitions over 10 k-folds.

ALS patients display the best results in terms of classification accuracy, and PD patients the worst results. Accordingly, it is worth mentioning that deviations of the ROC curves for PD patients are also much higher in comparison to other classes.

### 3.4 Confusion Matrix

The confusion matrix was built to depict the whole picture of the classifier’s performance: it allows seeing what percentage of each class was classified correctly, and where mistakes were made (Fig 6).

**Figure 6.**
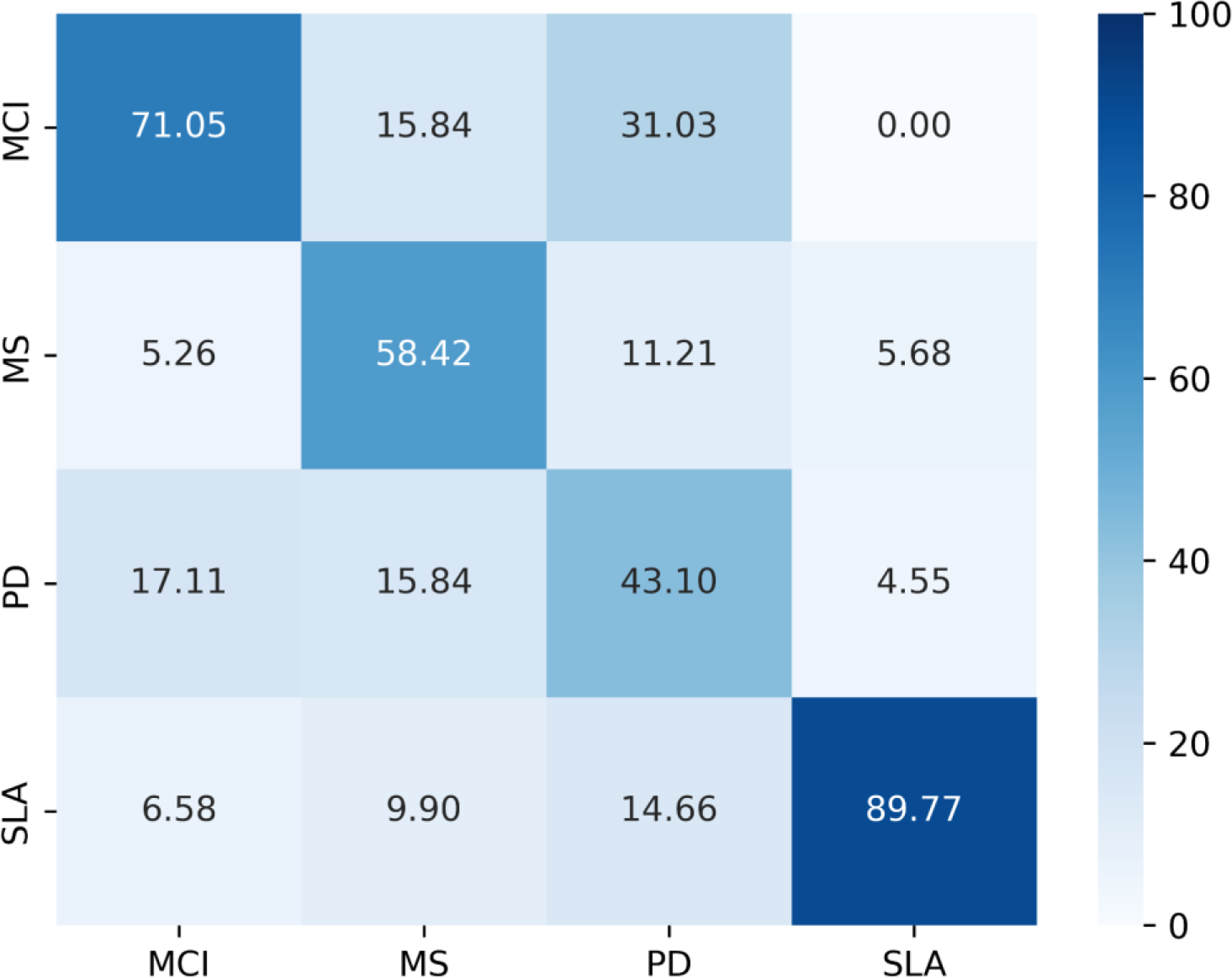
Confusion Matrix with relative percentage representation for 4 classes (SLA, PD, MS, MCI) with true values on y axis and predicted values on x axis for PLV features with LDA machine learning classifier.

Again, one can observe that the accuracy for PD patients is the worst, while the results for ALS subjects are the best. While these results might be affected by imbalanced classes, the results for all classes are well above chance level (which is equal to 35%).

### 3.5 Feature importance

The XGBoost classification algorithm allows us to quantify and compare the relative importance of the features during the classification process. To get valid results, we have defined 10 cross-validations with stratified KFolds. That is, the balance of the classes in the train and test splits are preserved. We run 40 times these stratified cross-validation iterations to reach convergence and then validate our results. After these steps, we obtained the feature importance for each of the features obtained from each FC metric taken separately (we focus here on the PLV, AEC, and ATM). Then, we evaluated and compared the features that showed the highest importance values for each of the sets considered (Fig. 7A)

**Figure 7.**
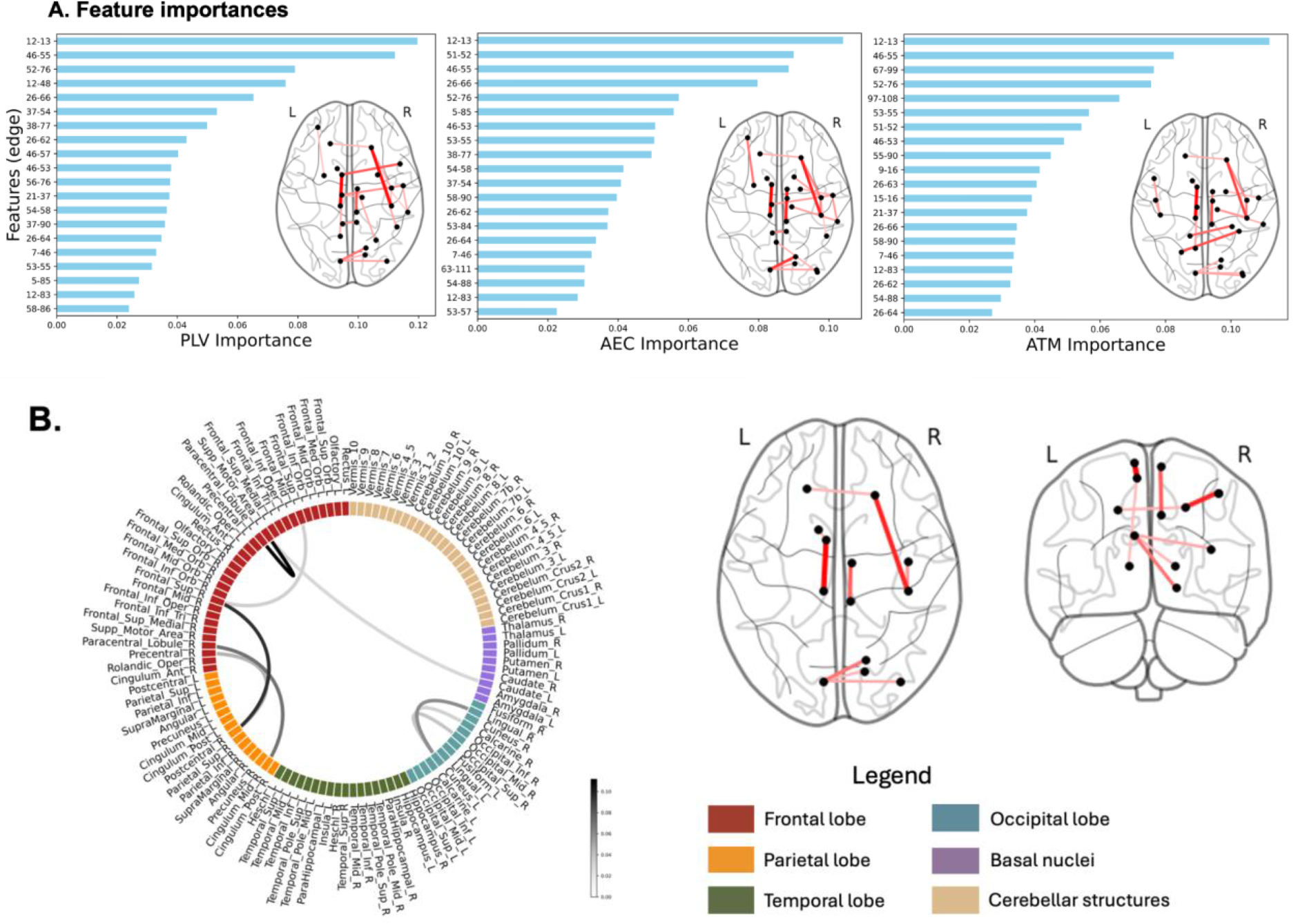
A. Feature importance for XGBoost across FC metrics along with these features on the brain plots. B. Connectome of overlapping features and its brain plot. The list of the edges is reported in the supplementary materials (see S6-S7-S8).

There are 9 overlapping features (i.e. edges) across 3 FC metrics (PLV, ATM, AEC), where we focused on the first 20 features with the largest feature importance according to XGBoost evaluation.

As an example of the consistency of our results across different metrics, the edge between the left supplementary motor area and the left paracentral lobule is the most important in all 3 feature sets. At the same time, the edge between the right frontal superior gyrus and right post central gyrus is also among the top 3 features across three feature sets.

Another interesting observation is that the left cuneus appears three times among the top pairs of edges (pairs of edges left cuneus and right lingual, left cuneus and Occipital middle gyrus, the left cuneus and the right calcarine cortex), meanwhile the left supplementary motor area, the right frontal superior gyrus and the right postcentral gyrus appear twice (for the exhaustive list of the region of interest see the supplementary material S3).

We selected 20 of the most significant features, according to their corrected pFDR values, and ranked them across feature sets (where rank 1 means the most significant, and rank 20 indicates the least significant feature). This way, we identified that edge features for the next pairs of ROIs: left supplementary motor area and left paracentral lobule, right frontal superior gyrus and right postcentral gyrus, right paracentral lobule and right middle cingulum are ranked as the top features (ranks 1 and 2) for three edge-specific FC metrics (AEC, PLV, ATM), therefore we see a strong overlap of features across metrics.

We noticed that 9 edges out of 20 selected were the same for 3 different metrics – AEC, PLV and ATM (Fig 7B)

To systematically test these findings, we applied pairwise Spearman correlation for the ranks of three metrics (Fig 8). This revealed that, indeed, PLV and AEC features’ ranks are highly positively correlated with a correlation coefficient equal to 0.89.

**Figure 8.**
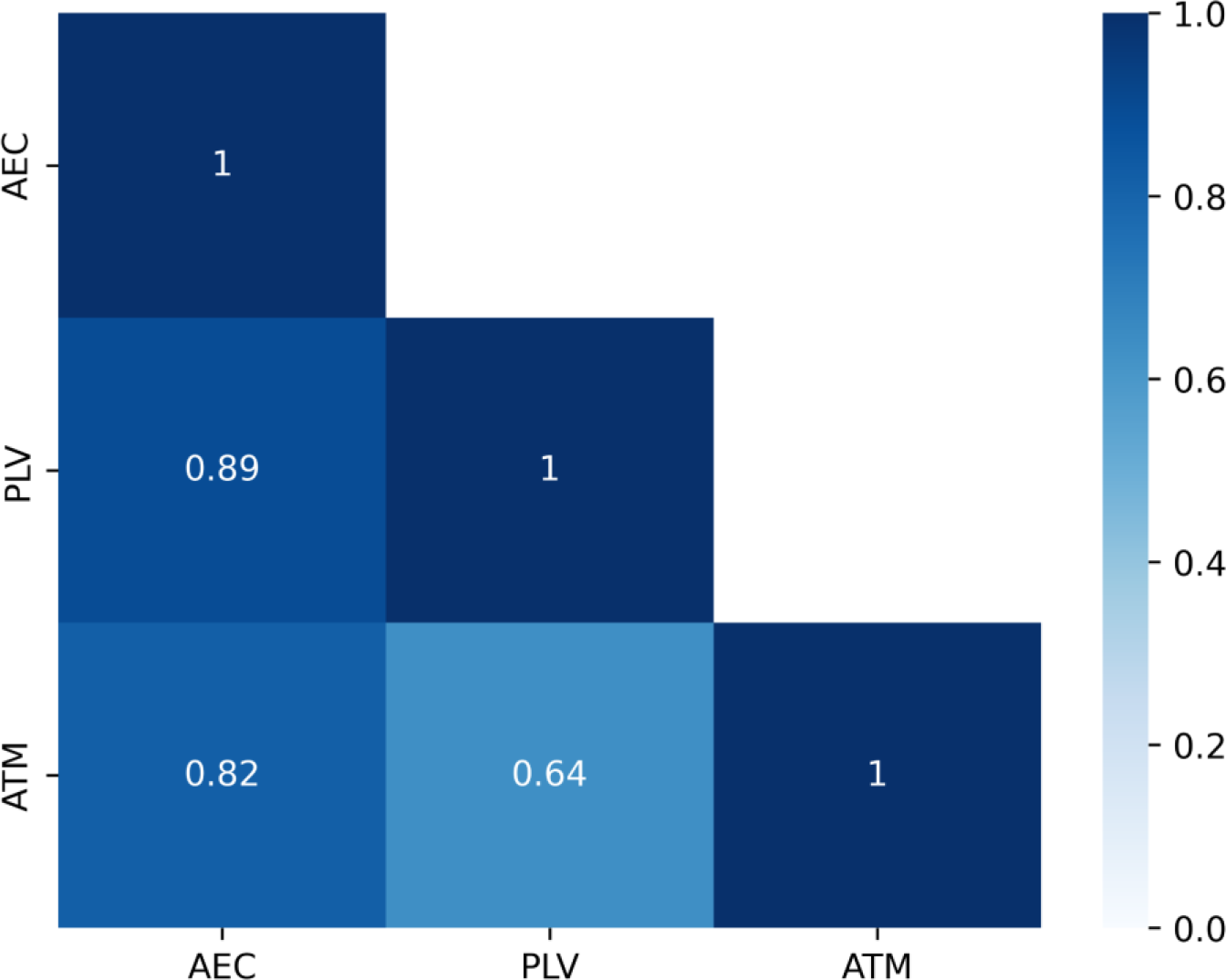
Feature importance for XGBoost across metrics.

We observed that AEC, PLV and ATM have 9 overlapping features out of the 20 best features according to the p-values, including the Cross Correlations leaves only one overlapping feature.

## 3. Discussion

In this study, we set out to identify a set of functional biomarkers to perform automated differential diagnosis (among MS, MCI, PD, and ALS) from MEG data. Our work focuses on the interpretability of the biomarkers, which is why we compared multiple connectivity metrics (AEC, PLV, Pearson’s correlation coefficient, and ATM) that are different in terms of interpretation. We tested the robustness of our analyses by feeding the data features to multiple classification algorithms (i.e., XGBoost, SVM, LDA). In particular, we used a vast amount of MEG data from a total number of 109 subjects affected by four different neurological diseases: ALS, PD, MCI, and MS. Firstly, from each cohort, we extracted different feature sets from four different FC metrics (PLV, ATM, AEC, and CC), each of which was obtained starting from a symmetric matrix, leading to a total number of 6670 edge-wise features. Due to the high dimensionality of our sample, we performed a Kruskal-Wallis test to reduce the dimensionality and to consider only the most discriminative features.

Firstly, our results showed more than 120 significant edge features among the four different feature sets (PLV, AEC, ATM, CC) for all the diseases. In particular, the Kruskal-Wallis test showed statistical significance (pFDR<0.0001) of edge-wise PLV values between the right frontal superior gyrus and the right postcentral gyrus. This finding might be related to the fact that the frontal lobe is involved in physiological processes related to motor function (which are notably impaired in SLA, PD, and MS) as well as to cognitive function, such as long-term memory (which is often impaired in MCI).

We then move on to estimate the accuracy of the selected features by adding the features in an iterative manner according to their p values. Such an approach enabled us to determine the optimal number of top features for each FC metric taken separately. The AEC reached a balanced accuracy of 63.07% with 38 features added, while the edge-wise PLV displayed the best-balanced accuracy (67,14%) with a total number of 34 added features. To our knowledge, no previous research has combined MEG data from patients with MCI, MS, PD, and ALS. However, some research has focused on these conditions individually. As an example, López ME et al. (López et al., 2014a) examined 105 subjects (36 controls and 69 MCI cases). They identified spectral bio-marked changes in the theta, alpha, and beta frequency bands in MEG data in MCI. Kim MJ et al. (M.-J. Kim et al., 2023) utilized a large EEG dataset including 417 MCI cases and applied a neural network that detected dementia with 81.1% accuracy. In the closely related work, Giovannetti A. et al. (Giovannetti et al., 2021) presented the Deep-MEG neural network, which was tested on 54 Alzheimer’s patients, each undergoing a five-minute resting state task. Similarly to our study, they used functional connectivity indices, phase locking values for classification. They reported an 87.4% AUC-ROC in identifying early MCI symptoms. For multiple sclerosis disease, using EEG, Kiiski H. et al. (Kiiski et al., 2018) assessed the responses of 35 subjects with multiple sclerosis during event-related potential cognitive tasks over three years. They found significant correlations between ERP visual components and cognitive function, identified using machine learning techniques. In related research, Karaca et al. employed a continuous wavelet transform to differentiate nine multiple sclerosis patients from 11 controls, achieving accuracy rates between 80%-88% in their best-performing models (Karaca et al., 2021). Furthermore, Ahmadi A. et al. analyzed five MS patients, developing a detection model using phase locking values and an online sequential extreme learning classifier, with performance scores ranging from 82% to 96% across different tasks (Ahmadi et al., 2019).

Our results are consistent across the three different ML algorithms (see Fig 3) with PLV and AEC showing the best accuracy with respect to ATM and CC. These results are in agreement with Chaturvedi et al., who showed that features extracted from the Phase lagIndex (PLI) were able to better discriminate between PD patients with and without MCI as compared to spectral features (Chaturvedi et al., 2019). These results might suggest that phase-based metrics might be more suitable in classification performance analysis as compared to amplitude-based metrics (i.e., power spectra). Phase-based metrics specifically capture synchronization among brain signals (defined as a bounded average phase difference). Synchronization is typically measured in the framework of the communication-through-coherence hypothesis (Fries, 2015), whereby communication among brain regions might be captured by the coherent activities of the corresponding brain signals. On the other hand, non-periodic activities, as well as simple correlation coefficients, seem to perform less well in this context. Regardless of the chosen FC metric, we see a consistent trend where the estimates at the edge level outperform those at the nodal level in terms of disease classification. On the one hand, nodal metrics capture the local activities and are predominantly sensitive to the dynamics of the local activations. On the other hand, edge metrics focus primarily on how brain regions interact among themselves. Therefore, our results might be interpreted as evidence that neurodegenerative diseases primarily alter how regions interact with each other at the large-scale level. In particular, since the PLV is the best-performing metric, this might be interpreted as the neurodegenerative diseases altering the ability of brain regions that are far apart to synchronize their activities.

While the PLV is sensitive to volume conduction, volume conduction does not offer a reasonable explanation for the ability to classify different subjects according to diagnosis. Furthermore, the edges of the ATM (that are more robust to volume conduction artifacts) also confirm the ability to correctly diagnose patients well above chance level.

Furthermore, the set of edges that contributed more to the classification were FC metric-independent. In general, it is interesting to note that the edges that are relevant to classification irrespective of the metrics are typically longer range connections, either in the antero-posterior direction of cross-hemispheric. Again, these results are representative of significant involvement and, as a consequence, impairment, of the frontal lobe in PD, ALS, MS and MCI respectively (Foong et al., 1997; Kendi et al., 2008; Trojsi et al., 2012; Wang et al., 2012).

Not many studies explain the rationale behind the feature extraction and selection method choice. It usually consists in a trade-off between enriching the information of interest and the risk of adding irrelevant inputs that could reduce the classification performance. Two types of approaches have been proposed. The first one consists in considering that fusing features will result in an improvement of the classification performance. For instance, Geraedts et al fused features obtained from the estimation of the power spectra in seven frequency bands (resulting in 16 674 features per EEG) before selecting them to discriminate cognitive functions in patients with Parkinson’s Disease during Deep Brain Stimulation (Geraedts et al., 2021). Similarly, López et al extracted spectral and non-linear metrics before fusing them and selecting them via their fast correlation-based filter to discriminate early Alzheimer’s disease and its prodromal form from healthy subjects (López et al., 2014b).

Another approach consists in fusing the classifiers’ output rather than the different types of features. Fusing the classifiers’ outputs confers a higher reliability and robustness through redundancy and facilitates the integration of heterogeneous data without normalizing them (Roli, 2009; Roli & Fumera, 2002; Ruta & Gabrys, 2000). In a recent work, we proposed a framework that was based on Riemannian geometry extended to functional connectivity measures through an ensemble learning method. We validated it on numerous publicly available datasets (Corsi et al., 2022). Such an approach notably ranked 1st in a clinical challenge that consisted in discriminating mental states from data obtained from stroke patients (Corsi et al., 2021). Future work will consist in considering this type of approach to enrich the information of interest used to discriminate diseases.

In conclusion, our is the first study investigating automated differential diagnosis in several neurological diseases, based on different connectivity metrics as well as on different classification algorithms. Our results demonstrate the existence of a common set of edges that drive the classification performance, irrespective of the particular metric chosen or the algorithm. These results demonstrate the existence of a robust set of long-range connections that are altered in neurodegeneration, across multiple diseases, and valid in terms of distinguishing specific diseases. Future studies will have to confirm the external validity of our results to different datasets and extend our analyses to more neurodegenerative diseases.

## Data Availability

All data produced in the present study are available upon reasonable request to the authors

## Data availability statement

The magnetoencephalography data and the reconstructed avalanches are available upon request to the corresponding author, conditional on appropriate ethics approval at the local site. The availability of the data was not previously included in the ethical approval, and therefore data cannot be shared directly. In case data are requested, the corresponding author will request an amendment to the local ethical committee.

## Competing interests

The authors report no competing interests

## Funding

This work was financially supported by Ministero Sviluppo Economico (Contratto di sviluppo industriale “Farmaceutica e Diagnostica” [CDS 000606]); European Union “NextGenerationEU,” (Investimento 3.1. M4. C2), project IR0000011, EBRAINS-Italy of PNRR and Contratto di sviluppo industriale-“Progetto CDS000904 -agevolazioni ex DM del 09/12/2014”

## Supplementary materials

**Supplementary materials 1:**
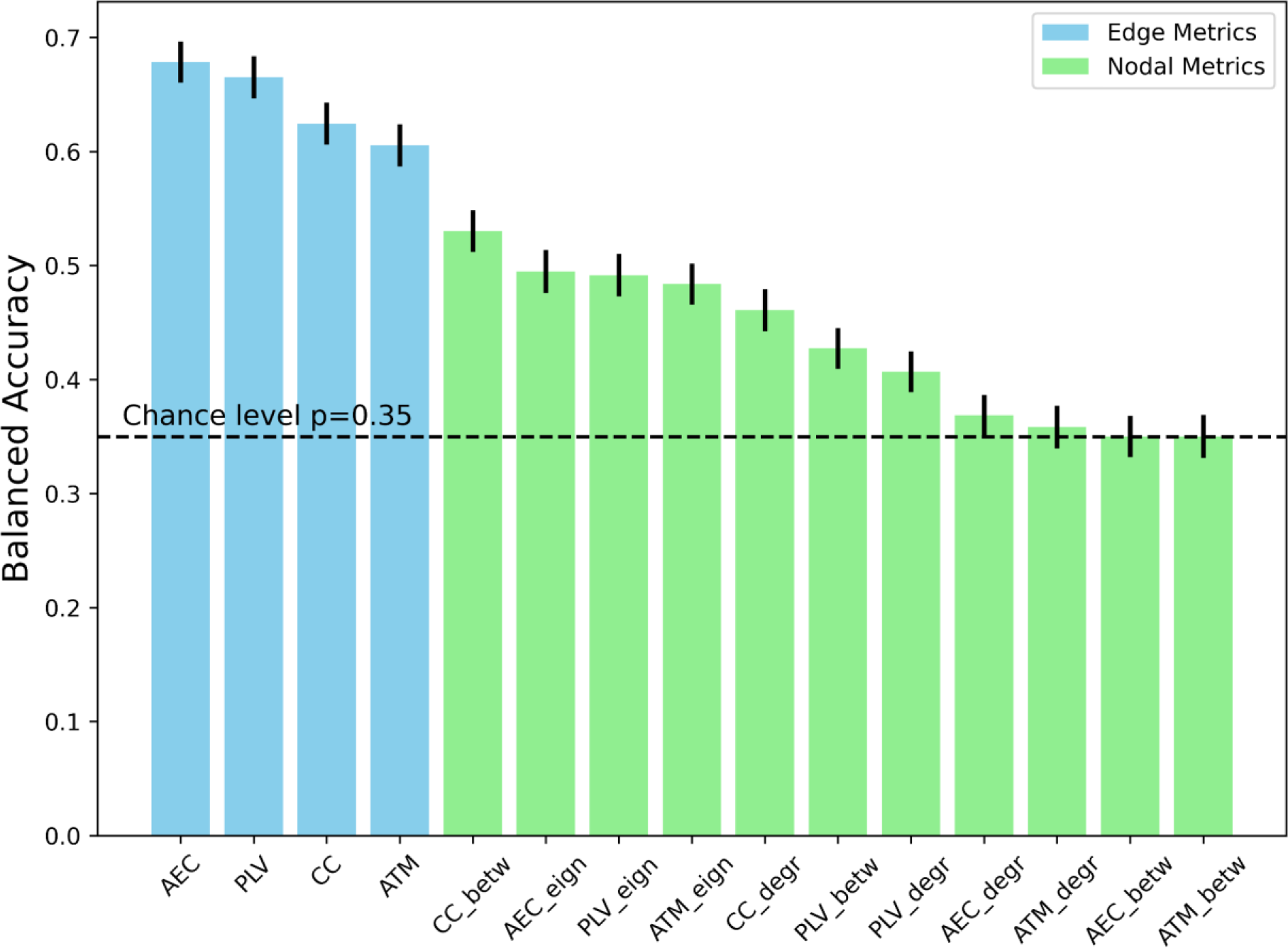
The balanced accuracies for all feature sets with SVM classifier. Each bar plot displays the averaged accuracy with its standard errors.

**Supplementary materials 2:**
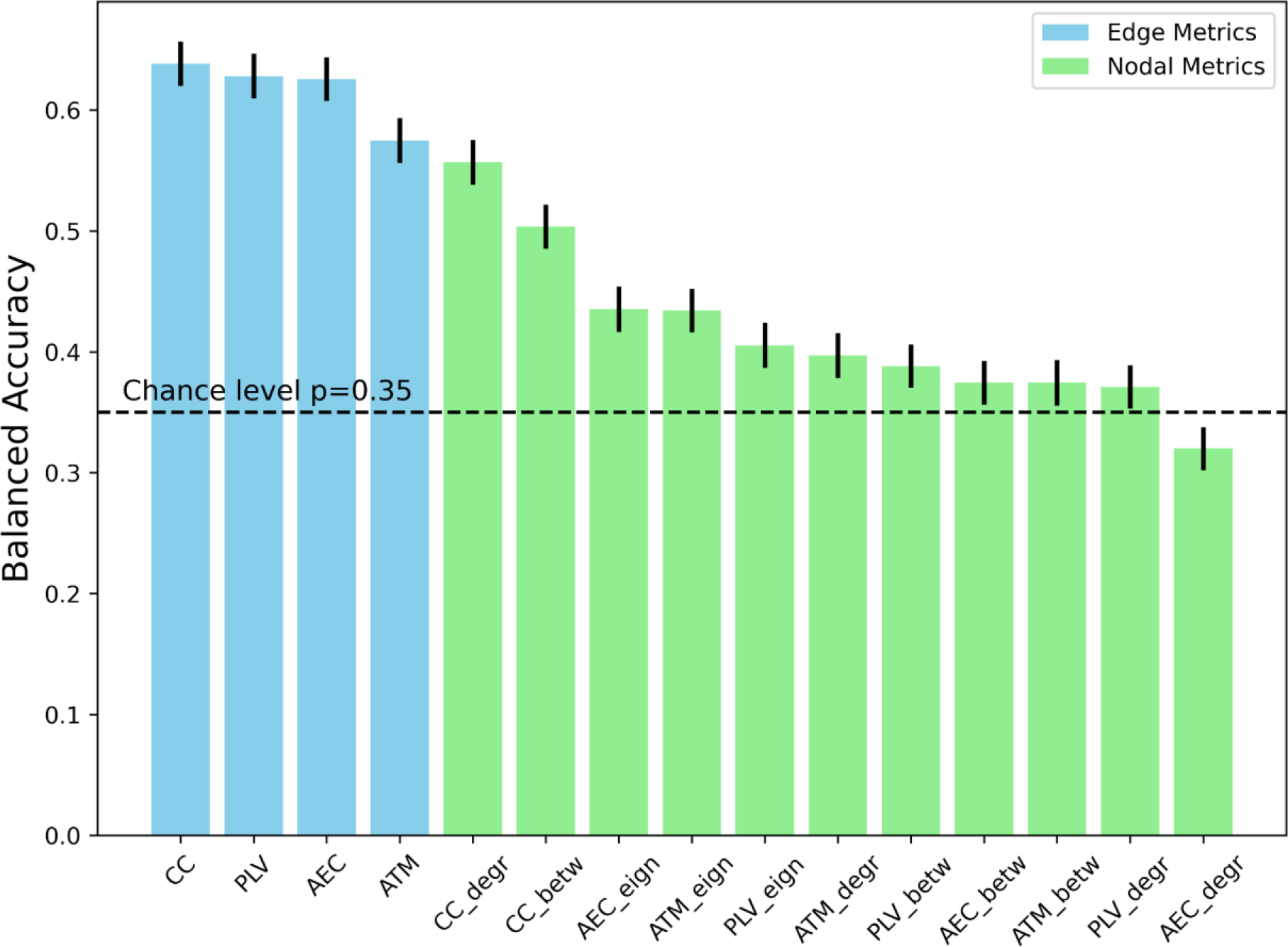
The balanced accuracies for all feature sets with XGBoost classifier. Each bar plot displays the averaged accuracy with its standard errors.

**Supplementary materials 3:**
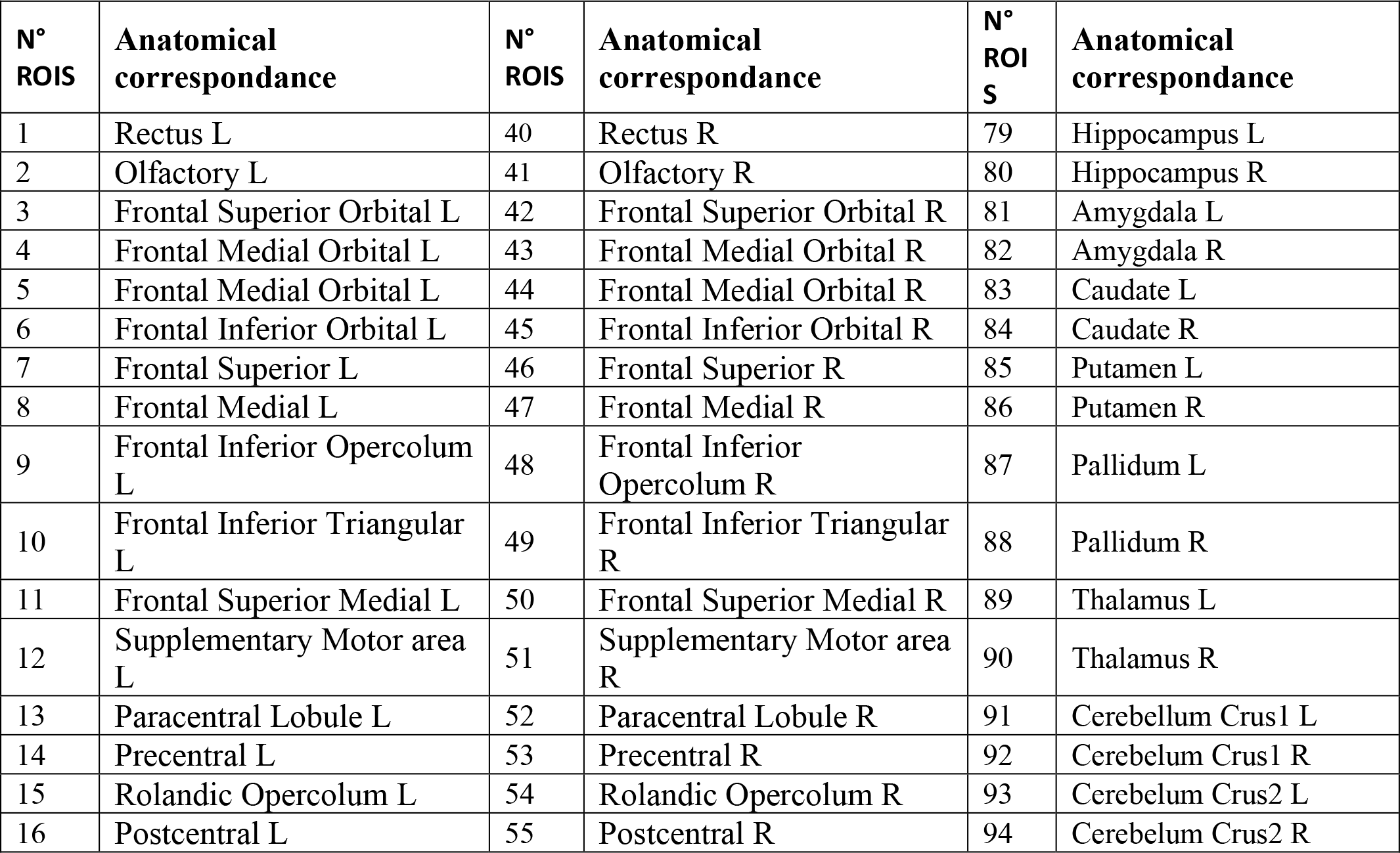
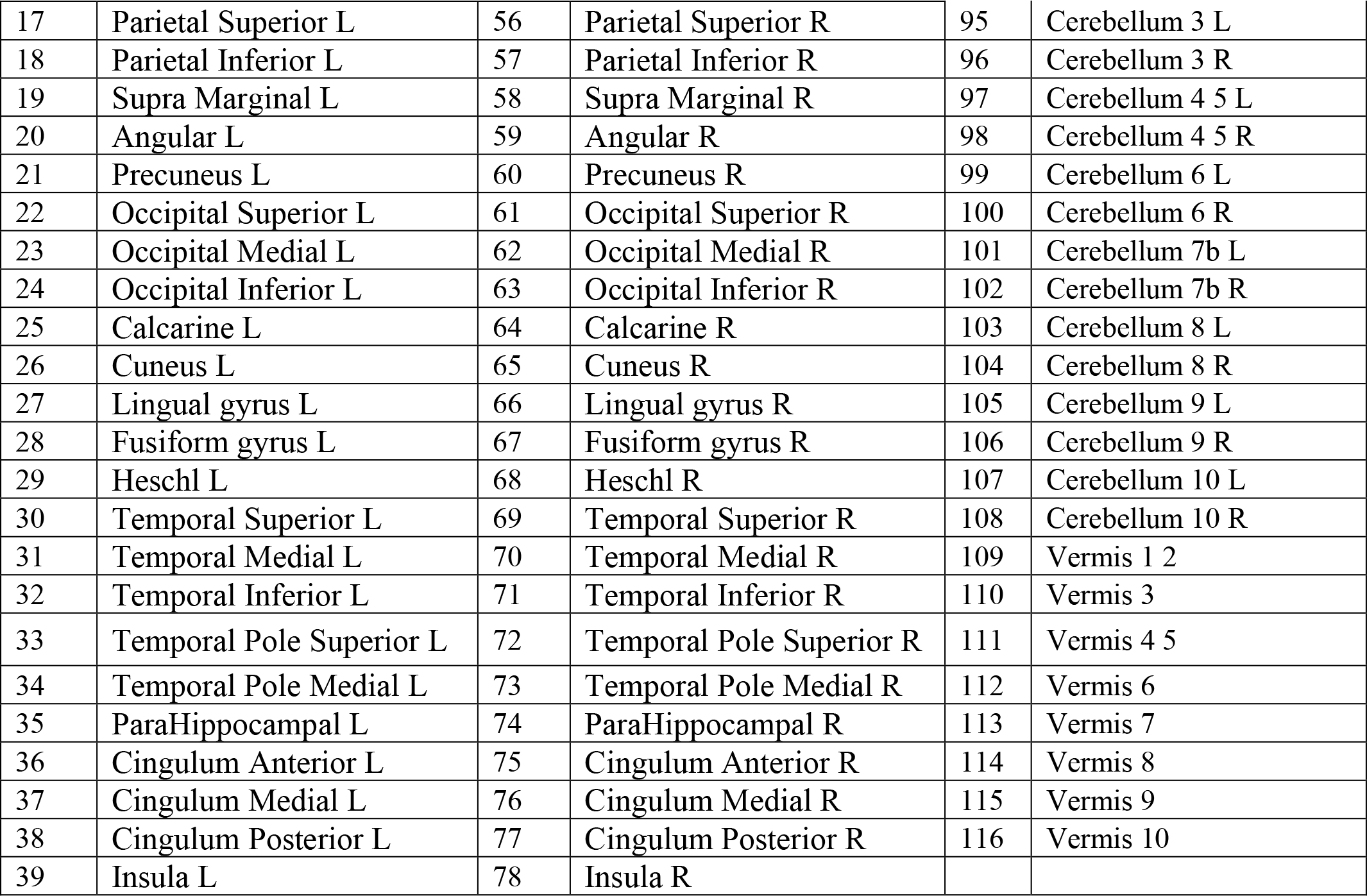
Exhaustive list of regions of interest.

**Supplementary materials 4:**
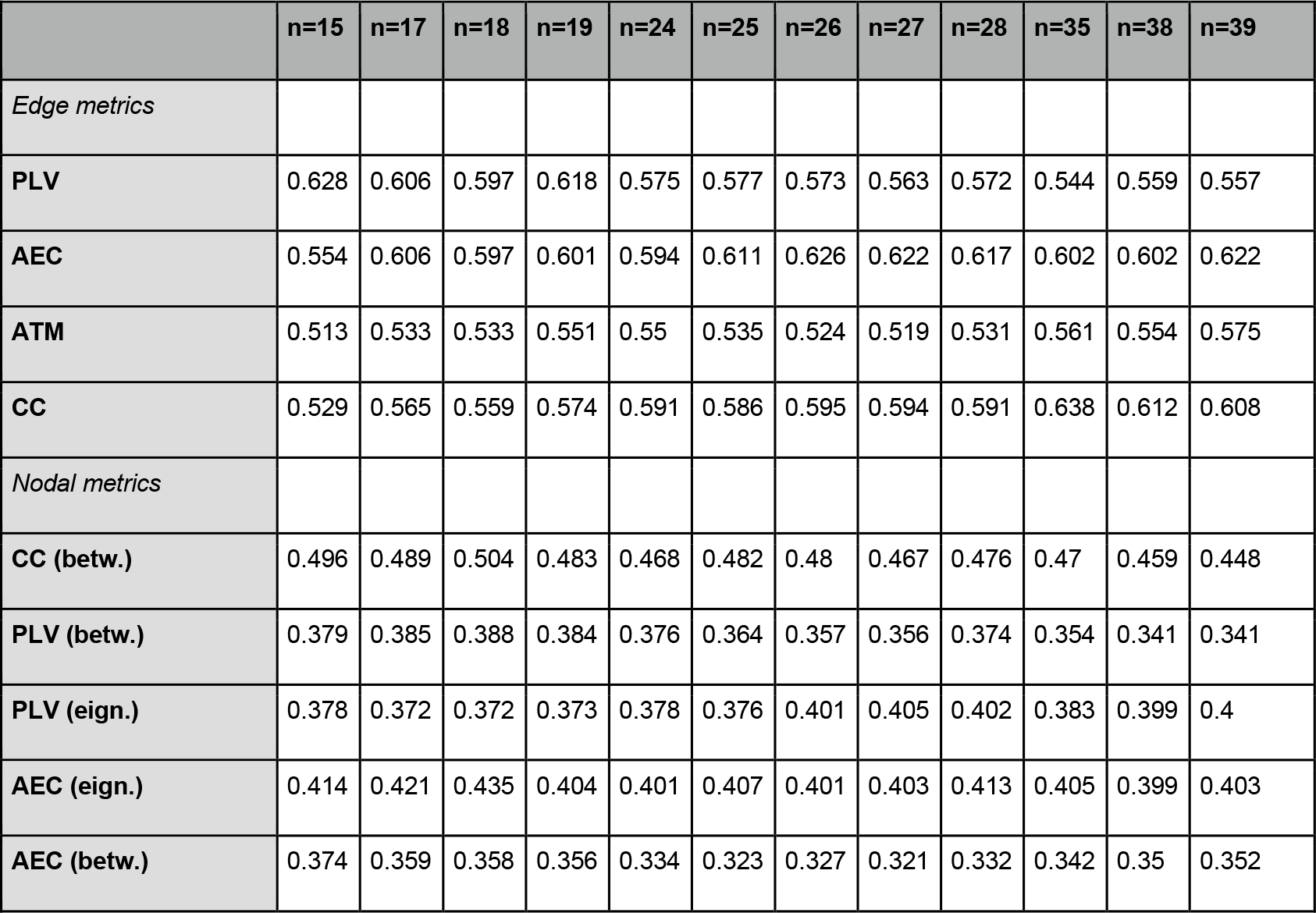
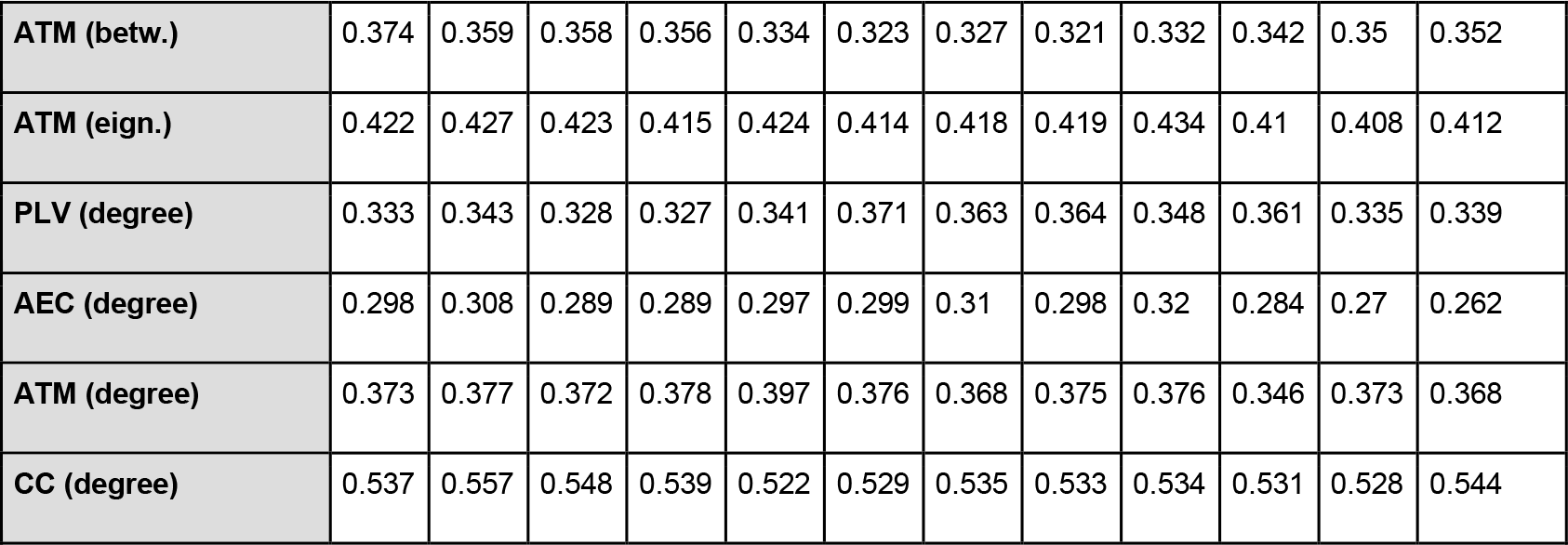
Balanced accuracy for the number of features which contain the best accuracy across different metrics (PLV, AEC, ATM, CC) with XGBoost.

**Supplementary materials 5:**
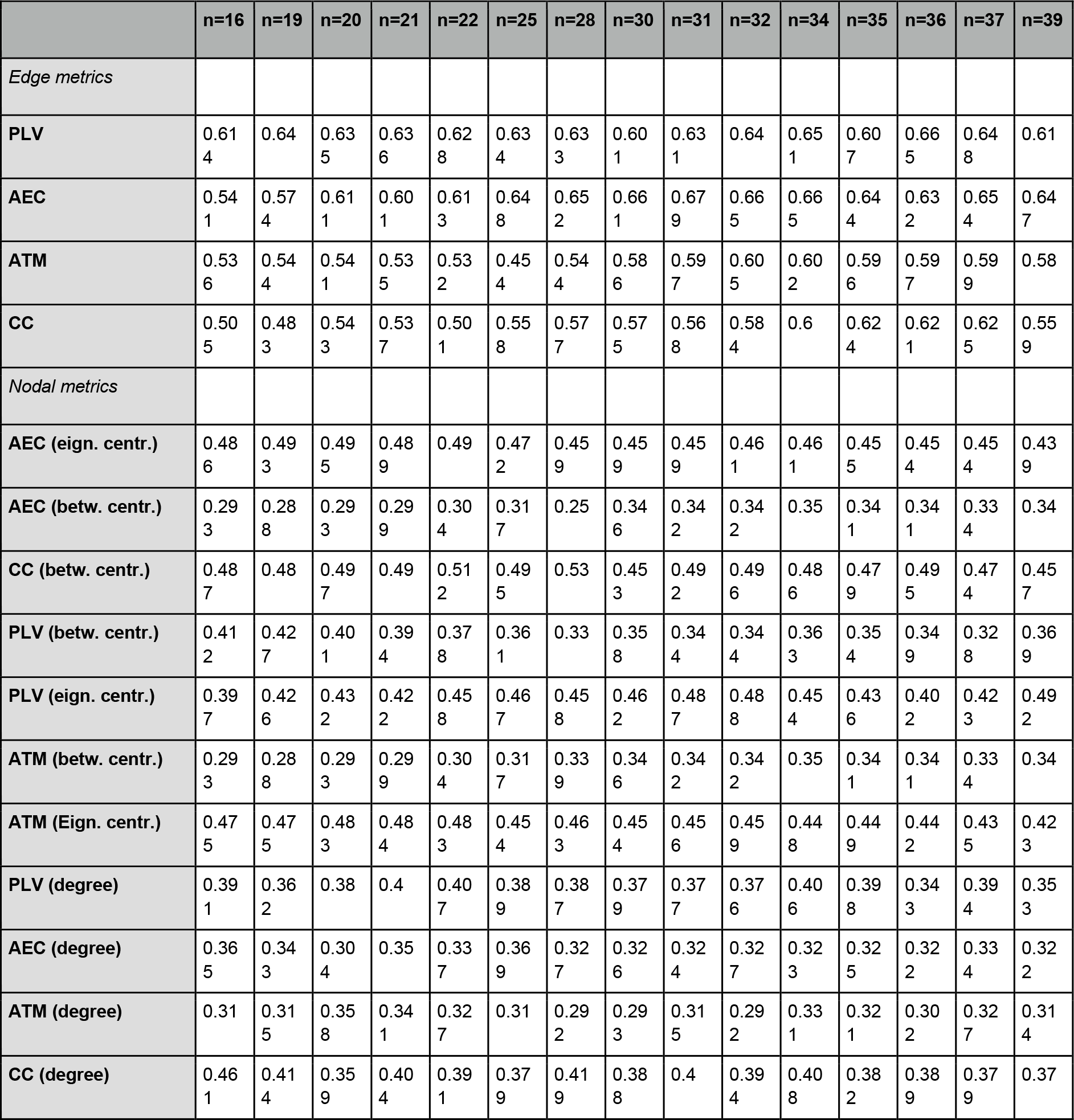
Balanced accuracy for the number of features which contain the best accuracy across different metrics (PLV, AEC, ATM, CC) with SVM.

**Supplementary materials 6:**
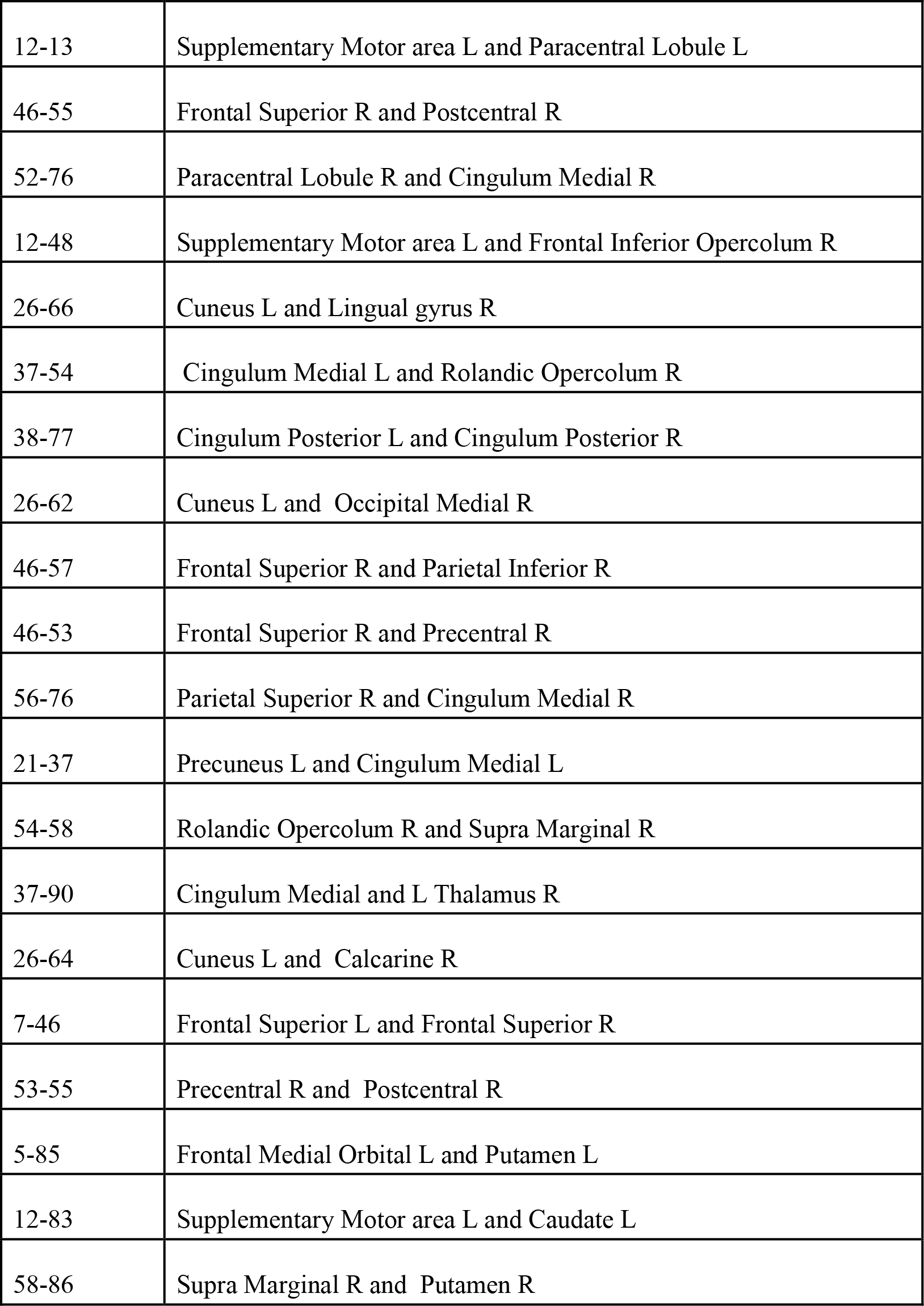
Features importance. List of associated edges for PLV.

**Supplementary materials 7:**
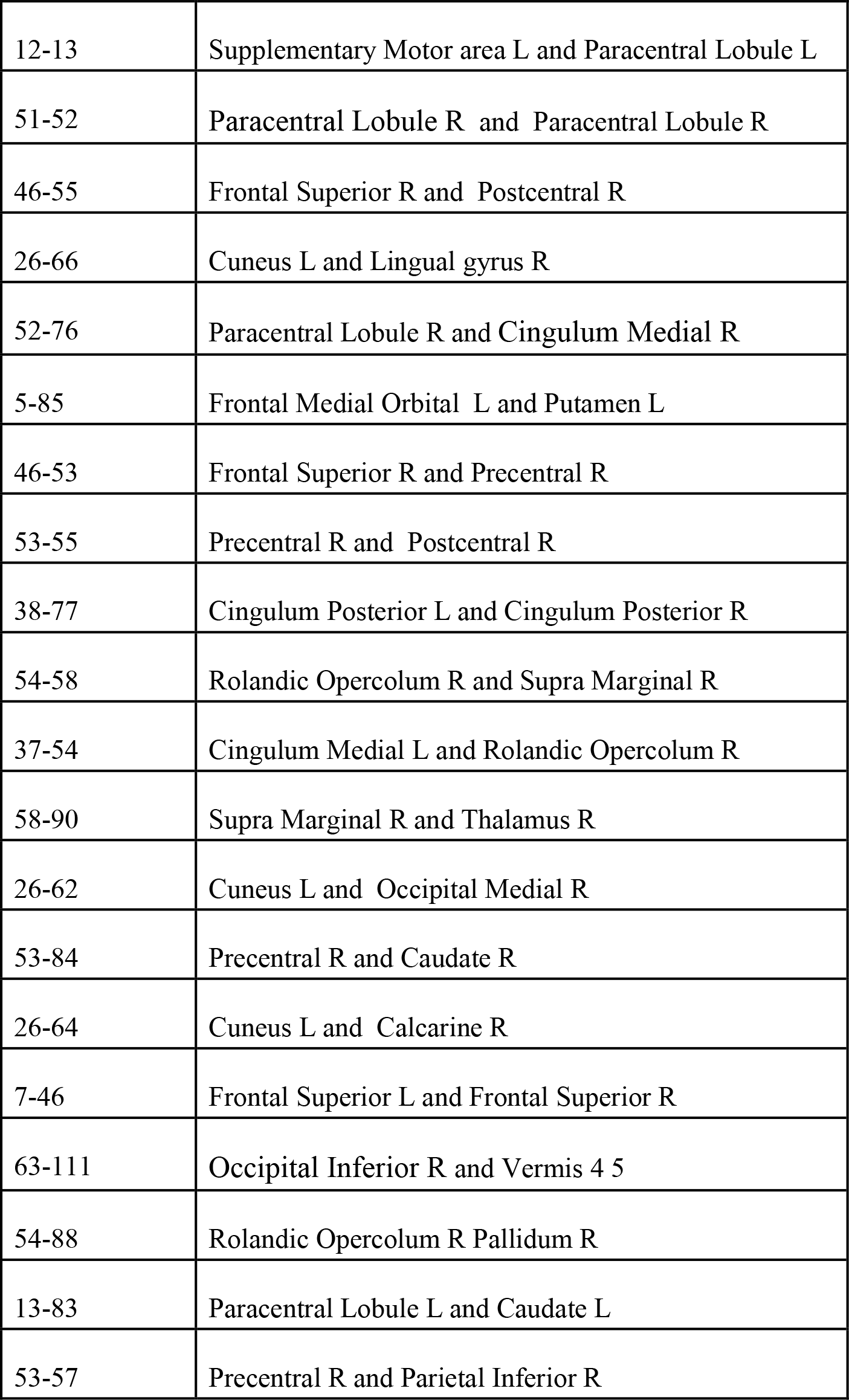
Features importance. List of associated edges for AEC.

**Supplementary materials 8:**
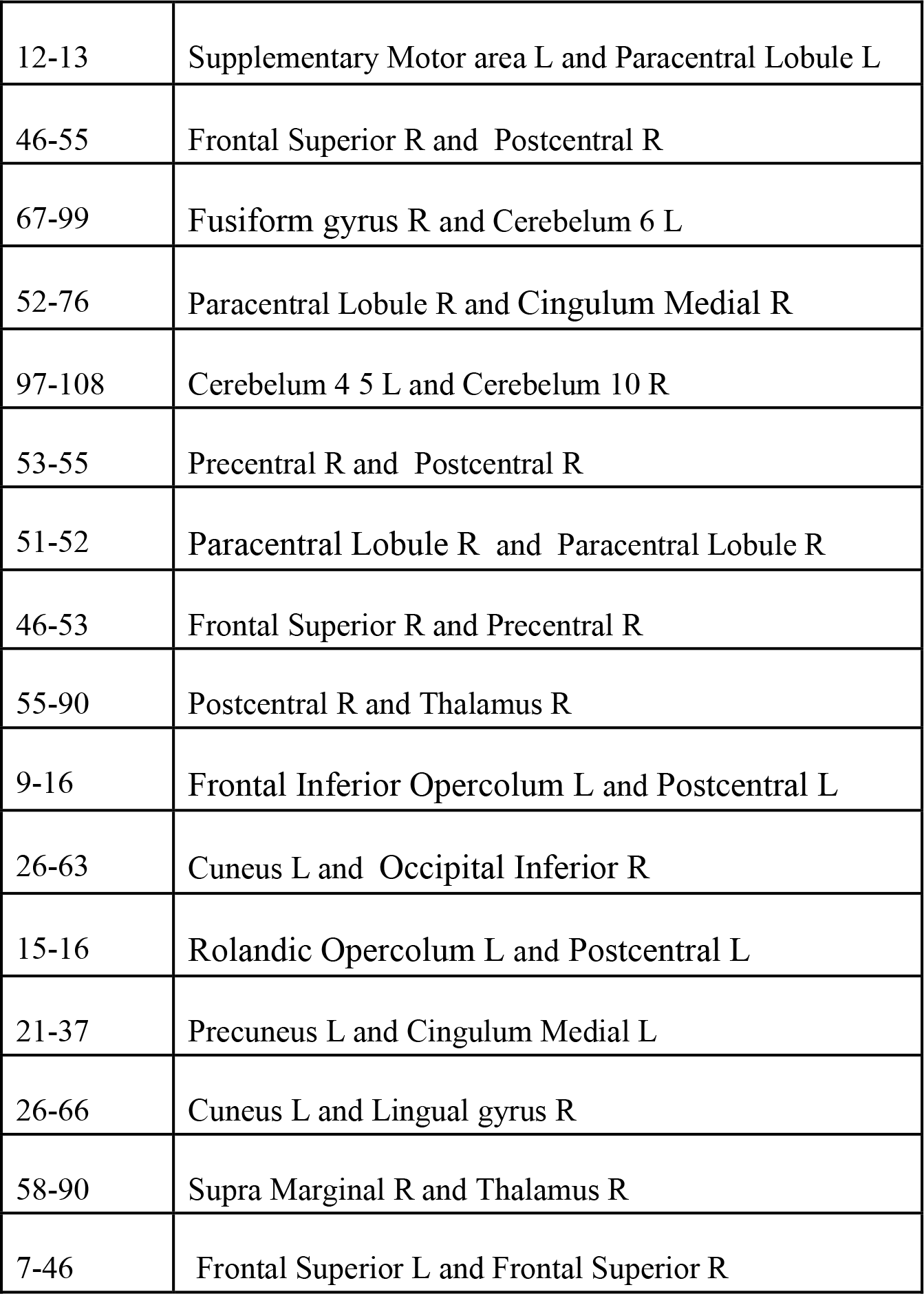
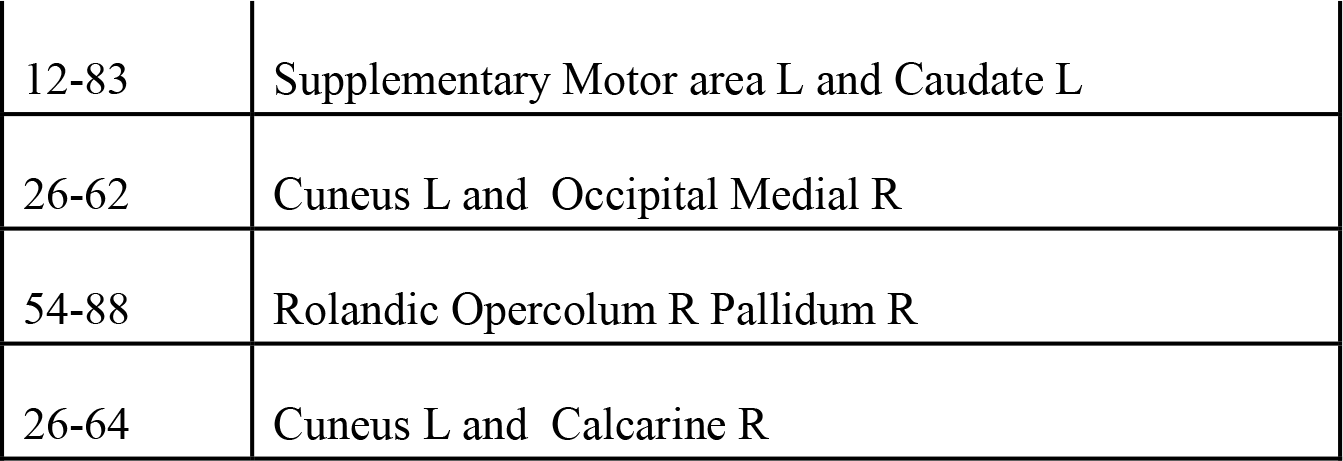
Features importance. List of associated edges for ATM.

## Notes

### Competing Interest Statement

The authors have declared no competing interest.

### Author Declarations

The study protocol was approved by the Comitato Etico Campania Centro (Prot.n.93C.E./Reg. n.14-17OSS)

